# Startle responses in Duchenne muscular dystrophy: a novel biomarker of brain dystrophin deficiency

**DOI:** 10.1101/2021.09.16.21263132

**Authors:** Kate Maresh, Andriani Papageorgiou, Deborah Ridout, Neil A. Harrison, William Mandy, David Skuse, Francesco Muntoni

## Abstract

Duchenne muscular dystrophy (DMD) is characterised by loss of dystrophin in muscle. Patients affected by DMD also have variable degree of intellectual disability and neurobehavioural co-morbidities. In contrast to muscle, in which a single full-length isoform (Dp427) is produced, multiple dystrophin isoforms are produced in the brain, and their deficiency accounts for the variability of CNS manifestations, with increased risk of comorbidities in patients carrying mutations affecting the 3’ end of gene, disrupting the shorter Dp140 and Dp71 isoforms. The *mdx* mouse model of DMD lacks Dp427 in muscle and CNS and exhibits exaggerated startle responses to threat, linked to the deficiency of dystrophin in limbic structures such as the amygdala, which normalise with postnatal brain dystrophin-restoration therapies. A pathological startle response is not a recognised feature of DMD, and its characterisation has implications for improved clinical management and translational research.

To investigate startle responses in DMD, we used a novel fear-conditioning task in an observational study of 56 males aged 7-12 years (31 DMD, mean age 9.7±1.8 years; 25 Controls, mean age 9.6±1.4 years). Trials of two neutral visual stimuli were presented to participants: one ‘safe’ cue presented alone; one ‘threat’ cue paired with an aversive noise to enable conditioning of physiological startle responses (skin conductance response, SCR; heart rate, HR). Retention of conditioned physiological responses was subsequently tested with presentation of both cues without the aversive noise in an ‘extinction’ phase. Primary outcomes were the magnitude of the initial unconditioned SCR and HR change responses to the aversive ‘threat’ and acquisition and retention of conditioned responses after conditioning. Secondary outcomes were neuropsychological measures and genotype associations.

The initial (unconditioned) mean SCR to threat was greater in DMD than Controls (Mean difference 3.0 µS (95% CI 1.0, 5.1), *P*=.004), associated with a significant threat-induced bradycardia only in the DMD group (mean difference -5.6 bpm (95% CI 0.51, 16.9); *P*=.04). DMD participants found the task more aversive than Controls, consequently early termination during the extinction phase occurred in 26% of the DMD group (vs. 0% Controls; *P*=.007).

This study provides the first evidence that boys with DMD show increased unconditioned startle responses to threat, similar to the *mdx* mouse phenotype that also responds to brain dystrophin restoration. Our study provides new insights into the neurobiology underlying the complex neuropsychiatric co-morbidities in DMD and defines an objective measure of this CNS phenotype, which will be valuable for future CNS-targeted dystrophin-restoration studies.

## Introduction

Duchenne muscular dystrophy (DMD) is an X-linked, life-limiting, progressive neuromuscular disorder with onset in early childhood, caused by mutations in the *DMD* gene encoding the protein dystrophin.^1^ The *DMD* gene is large (2.2Mb) and comprises 79 exons. It contains seven promoters for different dystrophin isoforms whose expression starts at different exons in the gene: three full-length 427kD isoforms (muscle, Dp427m; cerebral, Dp427c; Purkinje Dp427p, where 427 represents the molecular weight of the protein) and five shorter isoforms (Dp260, Dp140, Dp116, and Dp71, with its splice variant isoform Dp40), that exhibit developmental, regional and cell-type specificity within different tissues (Fig. 1).^2^ In the human brain, the full-length isoforms Dp427m and Dp427c, are present throughout the cortex and basal ganglia with the highest expression in the hippocampus and amygdala and lowest in the cerebellum, whereas Dp427p is expressed at lower levels. The shorter Dp140, Dp71 and Dp40 isoforms are also present in the brain: Dp140 is expressed at relatively high levels during brain development, with continued lower expression throughout adulthood^3^; Dp71 and Dp40 are alternative spliced products generated by the same promoter, with Dp71 being ubiquitously expressed in the brain including neurons, glia cells, blood vessels and the blood brain barrier.^4^ Dp260 and Dp116 isoforms are not expressed in the brain.^3^

**Figure 1.**
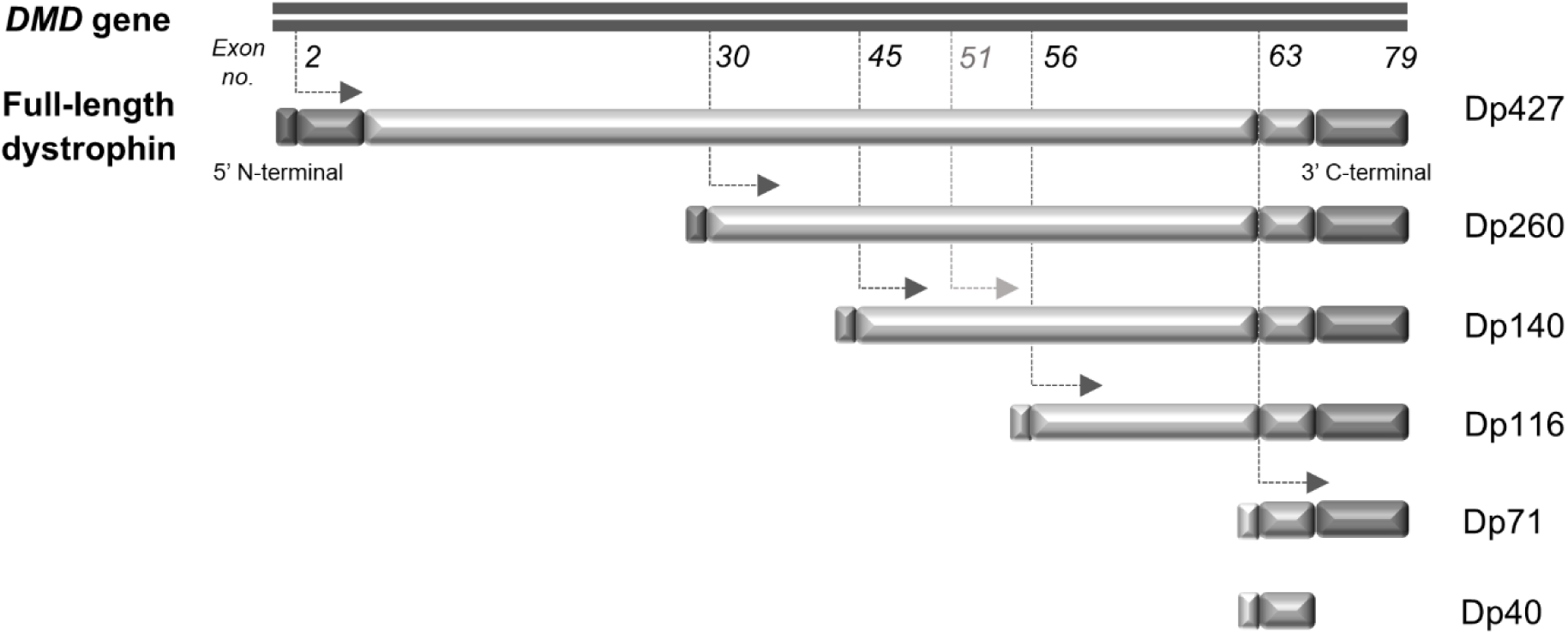
Schematic representation of *DMD* genomic organisation. The *DMD* gene, which encodes the protein dystrophin, is located on Xp21.2 and comprises 79 exons and seven promoters linked to unique first exons. Dotted arrows indicate the splice sites of these different internal promoters, which splice into the indicated exons (or preceding introns in the case of Dp140) to generate multiple dystrophin isoforms, shown in succession below the full-length dystrophin protein. The isoforms are named by their size in kiloDaltons (kDa). The three 427 kDa isoforms are Dp427m (muscle), Dp427c (cerebral), Dp427p (Purkinje); shorter isoforms are Dp260, Dp140, Dp116 and Dp71, the latter of which can be further alternatively spliced to form a 40 kDa Dp40 isoform. Each isoform has a different 5’ N-terminal domain, and all retain the same 3’ C-terminal domain. Numbers in italics indicate the exon number. In the case of the Dp140 isoform, transcription starts at intron 44 (just upstream of exon 45) but translation of the protein does not start until exon 51 (light grey arrow), leading to a long untranslated region (UTR) from intron 44-exon 50. It is difficult to predict the effect on Dp140 expression of mutations in this untranslated region. Adapted from figure in Muntoni, Torelli & Ferlini (2003)^2^

In all people with DMD, loss-of-function mutations in the *DMD* gene cause loss of the full-length Dp427 isoforms and lead to progressive skeletal muscle and cardiac dysfunction. The location of the *DMD* gene mutation determines whether expression of shorter dystrophin isoforms is preserved or also lost (Fig. 1). Of relevance to brain function, mutations upstream of exon 45 (towards 5’ end of the gene) do not affect expression of the Dp140 isoform (this group of patients is referred to as ‘Dp140+’) whilst those downstream of exon 50 lead to loss of Dp140 (termed ‘Dp140-’). Mutations between exons 45-50 have an indeterminate effect on Dp140 expression, due to a long untranslated region (UTR) encompassing these exons. Mutations towards the end of the *DMD* gene, between exon 63 and the 3’ C-terminal end, affect expression of the Dp71 isoform as well as the Dp140 isoform (termed ‘Dp140-/71-’).

DMD is associated with a complex neuropsychiatric phenotype, with approximately half of individuals with DMD being affected by problems including intellectual disability, language delay, autism spectrum disorder (ASD), attention-deficit-hyperactivity disorder (ADHD), emotional disorders and obsessive-compulsive disorder.^5,6^ Intellectual disability is strongly related to mutations disrupting the expression of Dp140 and Dp71 isoforms, with cumulative loss of these isoforms being associated with more severe intellectual impairment, with a mean IQ of 96 for boys only lacking Dp427, compared to 75 for boys also lacking shorter brain dystrophin isoforms (Dp140-/Dp71-).^7-9^ There is evidence that suggests there is also an increased risk of emotional, behavioural and neurodevelopmental disorders in individuals lacking the shorter dystrophin isoforms, but this is not confirmed.^10-12^ The heterogeneity of the neuropsychiatric presentation amongst individuals with DMD and the difference in phenotype in patients lacking different isoforms suggests that several pathophysiological mechanisms may be implicated in the different CNS phenotypes in DMD.

A mouse model of DMD, the *mdx* mouse, has a point mutation in exon 23 of the *DMD* gene,^13^ which abolishes expression of full-length Dp427 dystrophin without affecting any of the shorter isoforms. Preclinical studies in the *mdx* mouse have demonstrated impairments in long-term spatial and recognition memory, learning, cognitive flexibility and social communication.^14-17^ Increased fearfulness, stress reactivity and altered social behaviour are also seen, including enhanced defensive freezing behaviour during mild behavioural stress^18^ or foot shock^19^ in classical fear-conditioning paradigms. The parallels between the CNS phenotype in the *mdx* mouse and humans with *DMD* mutations suggest that much of this phenotype is due to loss of full-length Dp427 dystrophin.

In wild-type mice, Dp427 dystrophin co-localises with a subset of GABA_A_-receptors in the cerebral cortex, amygdala, hippocampus and cerebellum.^19^ GABA_A_-receptor distribution is decreased in *mdx* mouse amygdalae and a recent SPECT study also demonstrated a reduction of these receptors in the prefrontal cortex of adult humans with DMD.^20^ The Dp140 isoform is closely co-expressed with genes involved in early neurodevelopmental processes in humans,^3^ while murine studies have shown that Dp71 has a role in glutamatergic transmission and in glial-dependent extracellular ion homeostasis and Dp40 might play a crucial role in presynaptic functions in the brain.^21^

The reduced GABA_A_-receptor density in the hippocampus and amygdala is considered central to the pathological fear response phenotype seen in the *mdx* mouse.^22,23^ Postnatal dystrophin restoration in the brain of *mdx* mice is achieved using antisense oligonucleotides (AON), which induce the skipping of the affected in-frame exon 23, to cause an in-frame deletion that allows a shortened but functional dystrophin protein to be produced. Several studies have shown that such AONs can restore both dystrophin expression in the amygdala and postsynaptic GABA_A_-receptor density, and normalise the pathological fear response.^19,24^ Systemic AON therapies that induce exon-‘skipping’ to convert out-of-frame *DMD* mutations into ‘in-frame’ milder mutations that allow the production of shortened but functional proteins, are in current clinical trials or approved therapies in some countries.^25^ These AONs do not cross the blood brain barrier, although intrathecal administration would potentially allow brain dystrophin restoration.

These findings raise the intriguing possibility that AON therapies directed to restoring expression of brain dystrophin could also address some of the CNS comorbidities in DMD. Given the increasing life-expectancy in DMD with the current standards of care and emerging therapies, improving neuropsychiatric symptoms could significantly benefit quality of life. In order for any therapy to be evaluated, an outcome measure is needed to evaluate the effectiveness of dystrophin-restoration. The reversibility of the startle response in the *mdx* mouse after dystrophin-restoration therapy led us to propose that a similar phenomenon may occur in boys with DMD, but startle responses have not previously been investigated in DMD. Anxiety disorders are associated with increased startle responses,^26^ and anxiety symptoms are reported in 24-33% of people with DMD^11,27,28^ although this is a much less well-characterised feature than other CNS aspects of DMD.

Behavioural startle responses can be investigated using experimental fear-conditioning paradigms, where exposure to a threat stimulus causes an ‘innate’ or unconditioned physiological response. If a neutral stimulus is paired with the ‘threat’ stimulus, the physiological response becomes conditioned to occur with subsequent presentations of the neutral stimulus.^29^ We hypothesised that when tested using a fear-conditioning task, individuals with DMD would have heightened startle responses compared to control subjects. We designed a novel discrimination fear-conditioning task, which paired an aversive loud noise ‘threat’ stimulus with a neutral visual stimulus to elicit physiological startle responses that became conditioned to the neutral stimulus. These conditioned responses subsequently extinguished when the ‘threat’ was no longer presented in repeated trials.

Our aims were to determine whether group differences in physiological startle responses existed for: 1) initial unconditioned responses to threat; 2) acquisition of conditioned responses; 3) the degree of retention of conditioned responses when the neutral stimulus was presented without the ‘threat’, for DMD and Control groups. We also investigated associations between startle responses and trait anxiety and other neuropsychiatric symptoms, and differences between DMD genotype subgroups.

## Materials and methods

### Study design & setting

This observational cross-sectional study compared startle responses in young males with DMD with an age- and sex-matched control group using a fear-conditioning task. Study recruitment and testing took place from February to November 2019 at Great Ormond Street Hospital for Children (GOSH) and UCL Great Ormond Street Institute of Child Health.

### Participants

DMD and Control participants were recruited via: GOSH outpatient clinics (patients or siblings); a research database; advocacy groups; advertising to UCL staff and students. Eligibility criteria for all participants were: male, age 7-12 years, no significant visual or hearing impairment or noise-sensitivity. Specific criteria for DMD participants were a genetic diagnosis of DMD, not receiving ataluren (dystrophin-modulating therapy targeting nonsense mutations that may cross the blood-brain barrier); for Control participants no neurological or psychiatric diagnosis. All participants and parents/guardians were given age-appropriate written information about the study in advance, and informed written consent (parents) and assent (participants) was obtained according to the Declaration of Helsinki. All participants were allocated unique identifiers at recruitment. Ethical approval was granted by the Health Research Authority, London Bridge Research Ethics Committee (18.LO.1575).

Demographic and medical information was documented using a proforma: co-morbidities, medication, motor assessment, genotype information (DMD) and postcode (to estimate socio-economic status).^30^ Pubertal status was assessed using the ‘Growing and Changing’ questionnaire.^31^

### Task parameters

To determine physiological startle responses, we used a novel fear-conditioning task, described in detail in a previous methodology report^32^ and outlined in Figure 2. The task used neutral visual conditioned stimuli (CS-, the ‘safe’ cue and CS+, the ‘threat’ cue), which were different coloured squares presented on a laptop computer screen. An aversive auditory unconditioned stimulus (UCS; white noise approximately 85 dB) was presented binaurally through headphones. Four phases comprised: Pre-task calibration; Familiarisation (CS+ and CS-presented with no aversive UCS; eight trials); Acquisition (CS+ paired with UCS and CS-alone; three blocks of eight trials); Extinction (CS+ and CS-alone; five blocks of eight trials; occurred at least one hour after Acquisition). The CS+/CS-order was pseudo-randomised, and CS+ was presented with UCS in 10/12 CS+ trials (approximate 80% reinforcement schedule). CS colour was randomly counterbalanced amongst participants. UCS parameters: onset 5s after CS+ presentation, duration 1s, co-terminated with CS (CS duration 6s).

**Figure 2.**
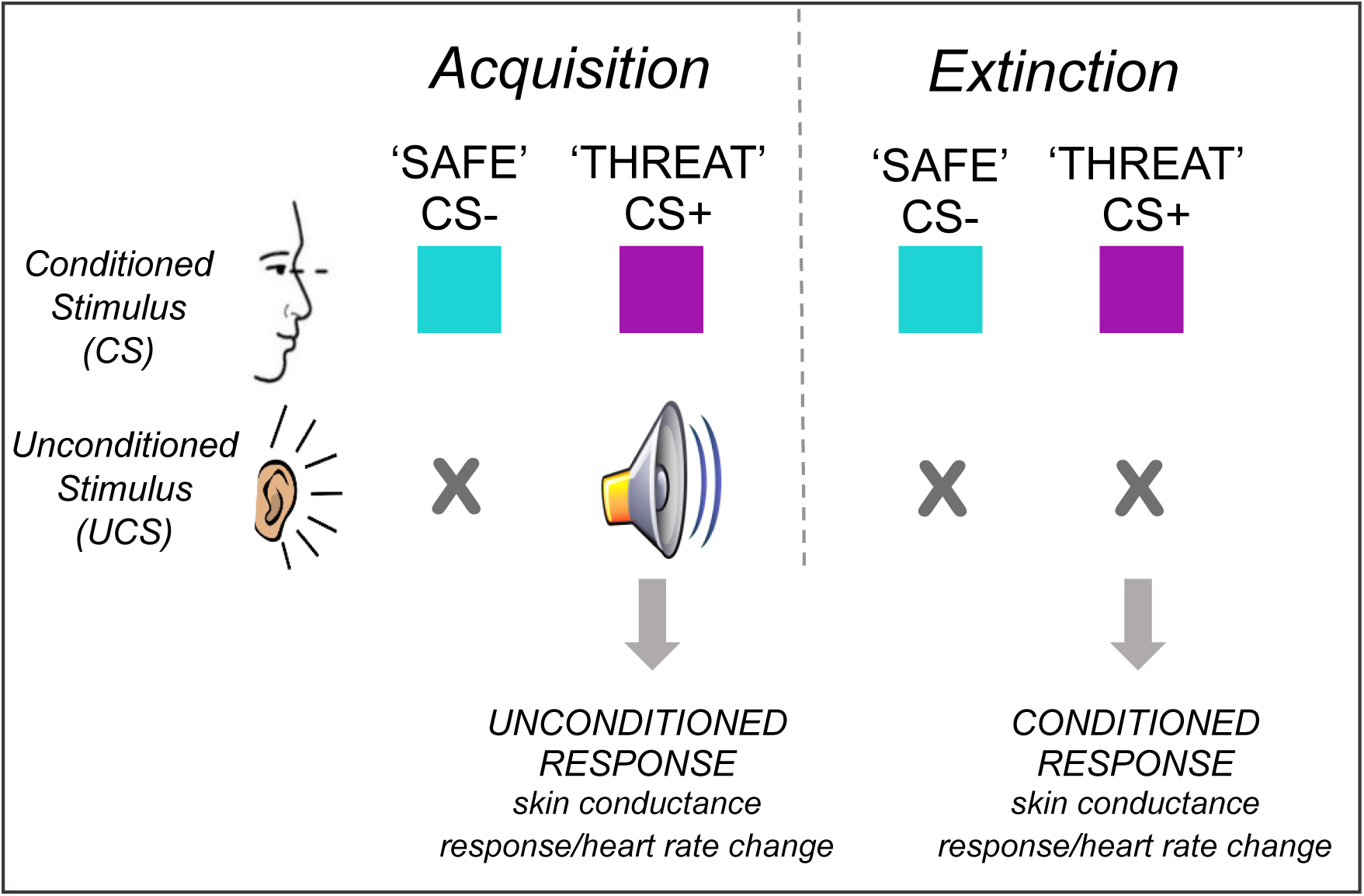
Schematic representation of the fear-conditioning task. Two different neutral stimuli, the conditioned stimuli (CS), are presented to the subject in randomised trials during the response acquisition phase (‘Acquisition’). One is a ‘threat’ cue (CS+) which is paired with an aversive noise stimulus, the unconditioned stimulus (UCS). A ‘safe’ cue (CS-) is presented alone with no UCS. At the initial presentation of the UCS, behavioural and physiological responses, termed ‘unconditioned responses’, are elicited. With repeated presentation of trials in the Acquisition phase, a learned association develops between the CS+ ‘threat’ cue and the unconditioned response, which then becomes ‘conditioned’. After 1-2 hours an Extinction phase is conducted, in which both CS+ and CS-cues are presented, but with no UCS. The CS+ cue initially elicits the conditioned response, but after repeated trials not reinforced by the UCS this association is extinguished and the CS+ cue no longer elicits the conditioned response, termed extinction. Prior to the Acquisition phase, a pre-task calibration phase and Familiarisation phase also occur, to allow physiological calibration manoeuvres, habituation to neutral stimuli and baseline measurements.

Skin conductance responses were obtained from electrodermal activity (EDA) recordings, measured using disposable surface skin electrodes on the non-dominant hand. Standard three-lead ECG recordings were obtained to determine heart rate. Physiological parameters were recorded using a Biopac MP160 unit with AcqKnowledge 5.0.3 software (Biopac Systems, Inc., Aero Camino, CA).

Direct neuropsychological assessment was conducted using the Wechsler Abbreviated Scale of Intelligence-II (WASI-II).^33^ This uses verbal and non-verbal reasoning tests to estimate intelligence quotient (IQ) without tasks that primarily involve working memory and executive function, which can be more impaired in boys with DMD and could confound general IQ estimation.^34^ Participants’ parent/carers completed neuropsychiatric symptom questionnaires: anxiety (Screen for Child Anxiety Related Emotional Disorders, SCARED)^35^; internalising and externalising problems (Child Behavior Checklist, CBCL)^36^; social communication problems (Social Communication Disorder Checklist, SCDC)^37^; inattention and hyperactivity (Conners’ Parent Rating Scale – Short version, CPRS-S).^38^ Several Control group scores were higher than published population mean scores (FSIQ-4 Mean difference 14.0, *P*<.001); self-report anxiety (*P*<.001); hyperactivity (*P*=.047) (Supplemental Table 1), therefore we also compared DMD group scores against age-/ gender-matched normative data for each scale.^33,36,38-40^

For DMD genotype analysis, subgroups were stratified by whether the genotype predicted expression of shorter dystrophin isoforms, similar to that described previously by Doorenweerd *et al*.^41^ The isoform outcome of mutation location was defined as follows: mutations upstream of intron 44 = Dp427 absent/Dp140 intact (termed ‘Dp140+’); intron 44 to exon 51 = indeterminate Dp140 expression (depending whether Dp140 promoter/long 5’UTR are affected; ‘Dp140_unk’); exon 51 to 62 = Dp140 absent (‘Dp140-’); exon 63 to 79 Dp427/Dp140/Dp71 absent (‘Dp140-/71-’).

A subgroup of DMD participants (n=11) repeated the fear-conditioning task after 3 months, confirming the test-retest reliability of the skin conductance response (results previously reported).^32^

### Data processing

EDA and ECG data were extracted from the 12s window post-CS onset, as determined from preliminary data from our group.^32^ Acquisition phase EDA data was also extracted for the 0-6s (First Interval Response, FIR) and 6-12s (Second Interval Response, SIR) windows to evaluate conditioned response acquisition and unconditioned response habituation respectively. EDA recordings represent the absolute skin conductance level (SCL) in microSiemens (μS), from which the skin conduction response (SCR) amplitude is derived: SCR = (SCL_max_) - (SCL_base_), where SCL_base_ is the baseline SCL at start of the trial, SCL_max_ is maximal SCL after CS onset. SCR metrics were defined for CS+ trials (SCR_CS+_), CS-trials (SCR_CS-_), Differential SCR, SCR_Diff_ = SCR_CS+_ - SCR_CS-_ in contiguous trials. All SCRs were visually inspected for artefacts by a blinded researcher, cross-referencing digital event markers. Where artefacts affected data, responses were re-measured, as detailed in a separate report^32^. For example, if SCL_base_ value was affected, amplitude was measured from the trough SCL prior to the response, SCL_min_, also derived during data extraction, or manually re-measured when SCL_min_ was not appropriate (for example if the minimum SCL occurred after the response peak). Where the SCR was completely obscured by artefact, these data were excluded. All exclusions were reviewed independently by another researcher. Non-responders were defined as participants in whom SCR was <0.01µS in the pre-task calibration manoeuvres and in ≥50% CS+/UCS trials, based on recommendations from previous literature.^42^

ECG data was transformed into R-R intervals (in seconds), the time interval between each heartbeat, where ‘R’ is the R-wave (maximal positive peak of the ECG complex). Heart rate (HR) HR = 60/(R-R interval) beats per minute (bpm). Heart rate in CS+ trials=HR_CS+_; heart rate in CS-trials=HR_CS-_. Change in HR (ΔHR) = (mean HR in sampling window) – (mean baseline HR), where baseline is mean HR in Familiarisation phase.

### Statistical considerations

The primary determinants of the sample size in this study (N=56, DMD n=31, Control n=25) were previous literature and pragmatic considerations rather than a formal power calculation, as an aim of this pilot was to gather data in this population for power calculations in future studies. However, based on previous healthy paediatric data we would expect sample sizes of n=28 per group (total N=56) to achieve a power of 80% to detect clinically important differences.^43^ Previous similar studies have examined fear responses in anxious/non-anxious children, with sample sizes of: 35 (subgroups n=17/18)^44^; 54 (subgroups n=16/38)^45^; 60 (subgroups n=8/11/19/16).^46^ This study was not designed to detect differences between the dystrophin isoform subgroups stratified by their Dp140 isoform expression profile, which were included as exploratory analyses only.

Analysis was conducted by a researcher blinded to participants’ group, with secondary ‘blind’ subject identifiers assigned by an independent researcher. All statistical analysis was performed with IBM SPSSv.26, using a two-tailed significance level, alpha, of *P*=.05 and, where appropriate, Bonferroni adjustment for multiple comparisons.

Plots of startle response metrics and scatterplots comparing variables were visually inspected to explore outcome distributions. Analysis of block data used linear mixed model analysis, to account for missing and unbalanced data, for i) SCR_Diff_ and ΔHR startle response metrics (fixed effects=Group (DMD, Control)/Block (FAM, ACQ1, ACQ2, ACQ3, EXT1, EXT2, EXT3, EXT4, EXT5); random effect=StudyID; Group*Block pairwise comparisons); ii) SCR_CS+_ and SCR_CS-_ in Extinction blocks (fixed effects=Group/Block/Stimulus (CS+/CS-); random effect= StudyID). Response extinction was defined as no significant discrimination between SCR_CS+_ and SCR_CS-_. The Residual Maximum Likelihood Estimation method was used in both analyses, with ‘Unstructured’ covariance structure.

Univariate analysis of variance was used to evaluate the unconditioned responses to the first reinforced CS+ trial (SCR_UC_ and ΔHR_UC_) and the conditioned SCR to the first CS+ trial in the Extinction phase (SCR_EXT_). These analyses included IQ as a covariate in the analysis, as there was evidence of IQ potentially being a confounding variable: IQ significantly differed between DMD and Control groups (*P*<.001), Dp140+ and Dp140-groups (*P*=.03), and when comparing DMD, Control and Dp140-groups with population normative data (Supplemental Table 1; P<.001). IQ also moderately correlated with SCR_UC_ in the Control group (*rho* 0.39, *P*=.06). Repeated measures analysis of variance was used for within group analysis of ΔHR_UC_. Two-tailed independent samples *t*-tests compared group means for neuropsychiatric scores. Spearman correlations were performed to investigate the association between neuropsychiatric scores and startle response metrics, and Chi-square tests compared task completion status between groups.

A sensitivity analysis was conducted on the primary outcome metrics to account for missing data, excluding ‘Non-completer’ participants who terminated the task early (completing <4 Extinction blocks), although all primary SCR and HR metrics were derived prior to this time point (Supplemental Table 6). The sensitivity analysis showed no change in statistical significance for all the primary SCR metrics (unconditioned and conditioned SCR_CS+_) and an increase in significance for ΔHR_UC_, therefore, the full dataset was used for the main analysis. A further sensitivity analysis was conducted on the HR metrics, excluding one DMD participant taking a B-blocker (which can lower heart rate); this showed no difference in outcomes (Supplemental Table 6).

## Data availability

The authors confirm that the data supporting the findings of this study are available within the article and its Supplemental material. In view of the rarity of DMD and that genotype data is included, only aggregated data will be made available on reasonable request.

## Results

### Participant demographics

Fifty-six participants took part in the study assessments (31 DMD and 25 controls), out of the 63 initially recruited (Supplemental Fig. 1). Mean group ages were: DMD 9.6 years (sd 1.4); Control 9.7 years (sd 1.8) (*P*=.80). There was no difference in mean pubertal stage or socio-economic status rank decile between the DMD and Control groups, nor between the DMD isoform subgroups (Table 1). In the DMD group, 29/31 were ambulant (mean NorthStar score 21.2/34), 30/31 were taking corticosteroid treatment and 7/31 were taking cardiac medication, as part of the DMD standards of care (7 taking Angiotensin Converting Enzyme inhibitors (ACEi), one of whom also took a Beta-blocker). Four DMD participants had previous diagnoses of ASD, and one of ADHD. Of the 56 physiological recording data sets, there were four exclusions from the SCR analysis: SCR responses were not recorded in two DMD participants due to technical equipment problems; one Control participant was excluded due to protocol deviation; one DMD participant was a ‘non-responder’ (1.9%, lower than the typically quoted SCR non-responder rate of 10%).^47^ Eight DMD participants requested early task termination (completing less than 4/5 Extinction block; termed ‘non-completers’), compared to no Control participants (*P*=.007).

**Table 1.** Participants’ baseline demographic information and available data for each group.

|  | Control | DMD | DMD Isoform Subgroups |  |  |  |
| --- | --- | --- | --- | --- | --- | --- |
|  |  |  | Dp140+ | Dp140- | Dp140-/71- | Dp140_unk |
| Demographics |  |  |  |  |  |  |
| No. participants | 25 | 31 | 12 | 11 | 1 | 7 |
| Mean age in years (std. deviation) | 9.7 (1.8) | 9.6 (1.4) | 9.6 (1.6) | 9.6 (1.5) | - | 9.8 (1.1) |
| Non-ambulant, n | - | 2 | 0 | 2 | 0 | 0 |
| Mean NorthStar score (0-34)<sup>a</sup> | - | 21.2 | 27.4 | 20.4 | - | 14.0 |
| Cardiac medication, n<br>(ACEi/beta-blocker)<sup>b</sup> | - | ACEi n=7;<br>BB n=1 | ACEi n=2;<br>BB n=1 | ACEi n=3;<br>BB n=0 | ACEi n=0;<br>BB n=0 | ACEi n=2;<br>BB n=0 |
| Corticosteroid treatment, n (Daily<br>(D)/Intermittent (I)/Alternate(A)) | - | 30<br>(D=10; I=19;<br>A=1) | 11<br>(D=8; I=2;<br>A=1) | 11<br>(D=6; I=5,<br>A=0) | 1<br>(I=1) | 7<br>(D=4; I=3;<br>A=0) |
| Autism spectrum disorder diagnosis | 0 | 4 | 0 | 4 | 0 | 0 |
| ADHD<sup>c</sup> diagnosis | 0 | 2 | 0 | 0 | 1 | 1 |
| Mean Socio-economic status rank<br>decile (1-10)<sup>d</sup> | 6.4 | 7.1 | 7.8 | 7.4 | - | 5.8 |
| Mean Pubertal Stage (1-5)<sup>e</sup> | 1.6 | 1.5 | 1.3 | 1.6 | - | 1.6 |
| Available data |  |  |  |  |  |  |
| Parent-report neuropsychiatric<br>scores, n | 25 | 31 | 12 | 11 | 1 | 7 |
| Participant self-report anxiety score,<br>n | 25 | 30 | 12 | 11 | 0 | 7 |
| Intelligence quotient assessment, n | 25 | 30 | 12 | 11 | 0 | 7 |
| Skin conductance response data, n | 24 | 28 | 10 | 11 | 1 | 6 |
| Heart rate data, n | 25 | 31 | 12 | 11 | 1 | 7 |
| Non-completers, n<sup>f</sup> | 0 | 8 | 2 | 5 | 1 | 0 |

<sup>a</sup> NorthStar score is a 17-item lower limb functional assessment scale used in DMD, scored out of 34.
<sup>b</sup> ACEi = Angiotensin Converting Enzyme inhibitor
<sup>c</sup> ADHD = attention deficit hyperactivity disorder
<sup>d</sup> Socio-economic status mean decile score was derived from the Index of Multiple Deprivation for England (Ministry of Housing, Communities & Local Government, 2019), available for participants living in England (n=52; DMD= 28, Control= 24).
<sup>e</sup> Pubertal stage was assessed using the parent/self-report 'Growing and Changing' questionnaire (Golding 1999). Scores range from 1 (pre-pubertal) to 5 (adult stage).
<sup>f</sup> Non-completers were participants who terminated the task early, completing <4/5 extinction blocks.

The eight DMD ‘non-completers’ comprised 2/12 Dp140+ participants, 5/11 Dp140-participants and 1/1 Dp140-/71-, showing a higher non-completion rate in those lacking Dp140, but this was not significant (*P*=.13) (Table 1). There were also no significant differences in the characteristics of the ‘Completer’ and ‘Non-completer’ groups (Supplemental Fig. 2; Supplemental Table 5). Data exclusion due to artefacts occurred in 3.8% of Control and 7.5% of DMD group Acquisition phase trials, rising to 16.5% in Control and 21.3% in DMD groups in Extinction phase trials (Table 1).

### Neuropsychological profile

DMD participants had an overall lower IQ than controls, and more symptoms of anxiety, inattention and other emotional and neurodevelopmental problems (Table 2). DMD participants scored 20-25 points lower than controls on all IQ measures, including Full-scale IQ, FSIQ: DMD 90.5 vs. Controls 115.4; (*t*=-6.7, *P*<.001). IQ measures were lower in the DMD subgroup lacking the Dp140 isoform (Dp140-) compared to those retaining it (Dp140+) (FSIQ Dp140+ 96.4 vs Dp140-82.8; *t*=-2.4, *P*=.03). The Control group had significantly higher FSIQ than the population normative score (mean difference 14.0, *t*=4.9, *P*<.001), therefore we also compared DMD group FSIQ against age-matched normative data (Supplemental Table 1).The Dp140+ subgroup mean FSIQ was not significantly different from the typical population (mean difference -5.0, 95% CI -14.0,4.0), *t*=-1.2, *P*=.25) whilst the Dp140-group had significantly lower FSIQ than the population norm (mean difference -18.6 (95% CI -27.2,-10.0), *t*=-4.8, *P*<.001). Due to the group differences in IQ and potential associations of IQ with primary outcomes, all subsequent analyses were adjusted for IQ.

**Table 2.** Neuropsychological assessment mean score comparisons between groups and subgroups.

| Neuropsychological assessment<sup>a</sup> | DMD vs. Control |  |  | DMD Dp140- vs. Dp140+ |  |  |
| --- | --- | --- | --- | --- | --- | --- |
|  | Raw score, mean difference (95% CI) | *t* | Sig., *P*<sup>b</sup> | Raw score, mean difference (95% CI) | *t* | Sig., *P*<sup>b</sup> |
| Full Scale Intelligence Quotient | -24.9<br>(-32.5, -17.4) | -6.7 | **<.001** | -13.6<br>(-25.3, -1.9) | -2.4 | **.03** |
| Anxiety | 5.4<br>(0.09, 10.8) | 2.0 | **.046** | 3.1<br>(-5.4, 11.5) | 0.8 | .46 |
| Internalising problems | 4.6<br>(1.2, 7.9) | 2.7 | **.009** | 4.3<br>(-1.5, 10.0) | 1.6 | .14 |
| Externalising problems | 6.5<br>(0.9, 12.2) | 2.3 | **.02** | 4.3<br>(-7.7, 16.3) | 0.7 | .47 |
| Social communication problems | 4.8<br>(1.9, 7.8) | 3.2 | **.003** | 4.4<br>(-0.9, 9.6) | 1.7 | .10 |
| Inattention | 2.0<br>(0.06, 4.0) | 2.1 | **.04** | 3.9<br>(0.6, 7.2) | 2.5 | **.02** |
| Hyperactivity | -0.12<br>(-2.3, 2.1) | -0.1 | .91 | 3.4<br>(0.2, 6.7) | 2.2 | **.04** |

<sup>a</sup> Full scale intelligence quotient (FSIQ-4) assessed using the Wechsler Abbreviated Intelligence Scale – 2nd Edition (WASI-II). The other neuropsychological/neuropsychiatric scores were obtained using parent-report questionnaires: Anxiety - Screen for Child Anxiety Related Disorders Parent-report (SCARED-P); Internalising and Externalising problems - Child Behavior Checklist (CBCL); Social communication problems - Social Communication Disorders Checklist (SCDC); Inattention and Hyperactivity - Conners' Parent Rating Scale-Revised, short version (CPRS-R).
<sup>b</sup> Between group comparisons performed with independent samples t-tests, showing mean difference and 95% confidence interval of the difference (95% CI). Sig. = Two-tailed significance, using alpha level of *P*=.05. *P*-values <.05 are highlighted in bold.

The scores for almost all neuropsychiatric co-morbidities were higher in the DMD group than Controls and population norms, including anxiety, internalising, externalising, social communication and inattention symptoms, and excepting only hyperactivity, although hyperactivity scores in both Control and DMD groups were significantly higher than the population normative scores (Table 2; Supplemental Table 1). Inattention and hyperactivity scores were significantly higher in Dp140-compared to Dp140+ subgroups, and the Dp140-subgroup scored significantly higher than the population mean in measures of anxiety, internalising problems, social communication problems, inattention and hyperactivity, whereas the Dp140+ subgroup had higher scores than the typical population only in internalising problems (Supplemental Table 1). In the whole cohort there were weak-moderate significant correlations (*rho*=0.3-0.4) between IQ and all neuropsychiatric symptom scores apart from hyperactivity (Supplemental Table 2), but not in the separate DMD and Control groups, apart from a significant negative correlation of inattention with IQ in the DMD group.

These data confirmed the divergence of the DMD participants from the Control group and normal population, in line with work from our group and others,^10-12^ with particular emphasis on the more pronounced cognitive and neuropsychiatric problems in the DMD subgroup lacking the Dp140 isoform.

### Physiological startle responses

#### Baseline physiological responses

Baseline mean SCRs did not differ between DMD and Control groups (mean difference 0.008µS (95% CI -0.18,0.20) *t*=0.09; *P*=.93) or Dp140+ vs. Dp140- (mean difference 0.08µS (95% CI -0.28,0.44), *t*=0.47; *P*=.64) (Fig. 3A). The DMD group had higher mean HR than the Control group (102.1bpm vs. 84.2bpm; mean difference 17.9bpm (95% CI 10.9,24.8), *t*=5.1, *P*<.001) (Fig. 3B), consistent with the well-recognised phenomenon of resting sinus tachycardia in DMD.^48^ There was no difference in baseline HR between DMD Dp140+ and Dp140-isoform subgroups (mean difference -7.7bpm (95% CI -19.0,3.7), *t*=-1.4, *P*=.18). Individual trial data is presented in Supplemental Fig. 3. Due to the potential confounding effect of IQ, physiological startle responses were adjusted for IQ (unadjusted data are available in Supplemental Table 4).

**Figure 3.**
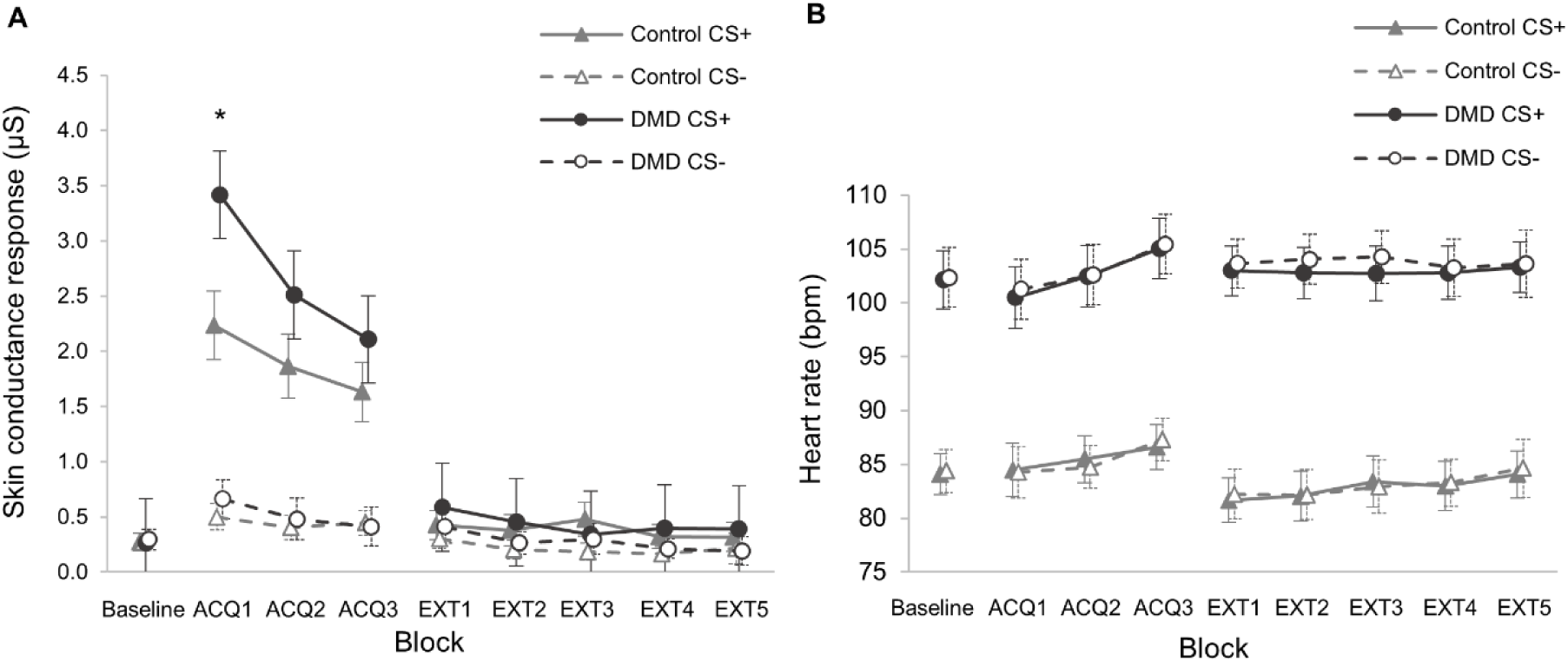
Mean physiological responses to ‘threat’ and ‘safe’ conditioned stimuli (CS+ and CS-) by block for all fear conditioning task phases in DMD and Control groups. (**A**) Skin conductance responses (SCR, measured in microSiemens, µS). Mean SCR in first acquisition block (ACQ1) was significantly higher in the DMD compared to control group (\**P*=.03). (**B**) Heart rate (HR, measured in beats per minute, bpm). SCR derived from electrodermal activity (EDA) recorded from the palmar surfaces of digits 2 & 3, and defined as the baseline-to-peak EDA in the 12s window following CS presentation. HR derived from the inter-beat interval from 3-lead electrocardiogram (ECG) recording. Error bars show the 95% Confidence Interval.

#### Increased unconditioned startle responses in DMD group

The mean unconditioned SCR to the initial threat stimulus (SCR_UC_) was significantly higher in the DMD group compared to Controls, with the greatest difference on the first CS+ trial (mean difference 3.0µS (95% CI 1.0,5.1), *P*=.004)) (Table 3; Fig. 4), although mean SCR was also higher in the DMD group compared to Controls for the whole first Acquisition block (mean difference 2.2µS (95% CI 0.9,3.5), *P*=.001; Fig. 3A).

**Table 3.** Primary outcomes: skin conductance and heart rate response metrics in DMD group and DMD isoform subgroups compared to Control group.

|  | DMD vs. Control |  |  | DMD Dp140+ vs. Control |  |  | DMD Dp140- vs. Control |  |  | DMD Dp140- vs. Dp140+ |  |  |
| --- | --- | --- | --- | --- | --- | --- | --- | --- | --- | --- | --- | --- |
| Outcome measure | Mean diff. (95% CI) | *F* | Sig., *P*<sup>d</sup> | Mean diff. (95% CI) | *F* | Sig., *P*<sup>d</sup> | Mean diff. (95% CI) | *F* | Sig., *P*<sup>d</sup> | Mean diff. (95% CI) | *F* | Sig., *P*<sup>d</sup> |
| $SCR_{UC}$ (in μS)<sup>a</sup> | 3.0<br>(1.0, 5.1) | 8.9 | **.004** | 3.4<br>(1.3, 5.6) | 10.4 | **.003** | 3.9<br>(1.1, 6.6) | 7.8 | **.008** | 0.61<br>(-3.0, 4.2) | 0.1 | .73 |
| $\Delta HR_{UC}$ (in bpm)<sup>b</sup> | -8.7<br>(-16.9, -0.51) | 4.5 | **.04** | -10.0<br>(-19.8, -0.19) | 4.3 | **.046** | -4.2<br>(-17.6, 9.1) | 0.4 | 0.53 | 3.5<br>(-6.3, 13.2) | 0.6 | .47 |
| $SCR_{EXT}$ (in μS)<sup>c</sup> | 0.37<br>(-0.23, 0.96) | 1.5 | .22 | 0.34<br>(-0.21, 0.89) | 1.6 | 0.22 | -1.1<br>(-2.1, -0.21) | 6.2 | **0.02** | 0.84<br>(-0.11, 1.8) | 3.6 | .08 |

<sup>a</sup> $SCR_{UC}$ = Unconditioned skin conductance response to the first ‘threat’ trial with conditioned stimulus, CS+, in microSiemens ( μS).
<sup>b</sup> $\Delta HR_{UC}$ = Unconditioned change in heart rate response to the first CS+ ‘threat’ trial, in beats per minute (bpm).
<sup>c</sup> $SCR_{EXT}$ = Conditioned skin conductance response to the first CS+ trial of the Extinction phase, in μS.
<sup>d</sup> Significance (Sig.) testing using univariate analysis of variance between groups/subgroups, adjusted for Full Scale IQ (FSIQ), taking alpha of $P=.05$ ; $P$ -values <.05 are highlighted in bold. $F$ = effect size of Group/Isoform group. Bonferroni adjustment made for multiple comparisons. Covariate values for FSIQ=101.9 for DMD vs. Control; FSIQ=109.5 for Dp140+ vs. Control; FSIQ=104.8 for Dp140- vs. Control; FSIQ=89.3 for Dp140- vs. Dp140+). FSIQ data available for 30/31 DMD, 25/25 Control, 12/12 Dp140+ and 11/11 Dp140- participants.

**Figure 4.**
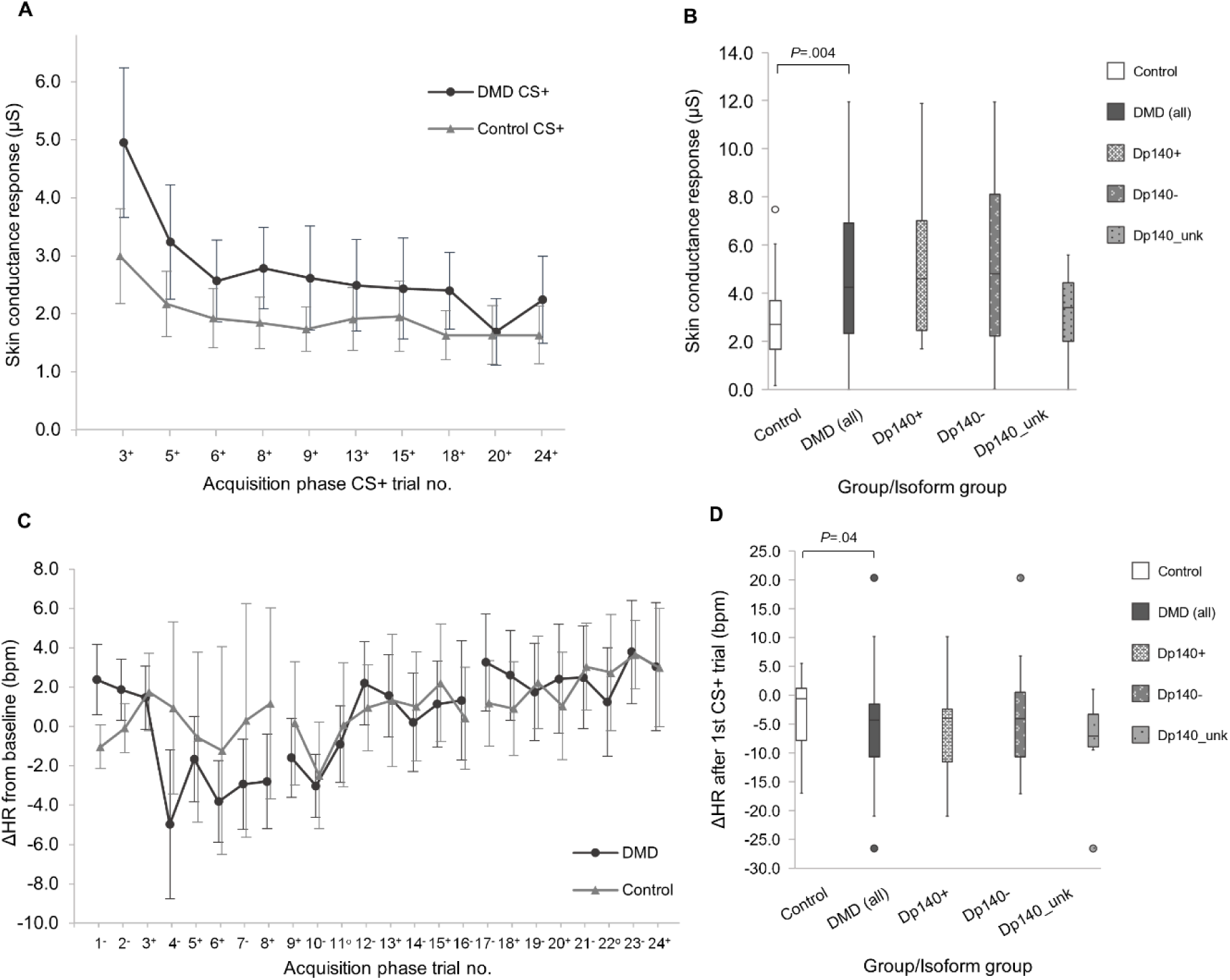
Unconditioned responses to the aversive threat stimulus. (**A**) SCR_CS+_ for DMD and Control groups in Acquisition phase. Two ‘threat’ conditioned stimulus (CS+) trials were omitted (Acquisition trial 11 and 22) as these were unreinforced CS+ trials (no aversive noise presented). (**B**) Box plot of unconditioned SCR_CS+_ for the first CS+ trial (SCR_UC_) for Control, DMD, and DMD isoform subgroups categorised by Dp140 isoform status. The Dp140+ group comprises DMD participants who retain the Dp140 isoform (n=10); Dp140-group comprises DMD participants who lack the Dp140-isoform (including Dp140- and Dp140-/71-; n=12); Dp140_unk group is DMD participants whose Dp140 status is uncertain (n=6). (**C**) Change in heart rate (ΔHR) from baseline in beats per minute (bpm) for DMD and Control group for all Acquisition phase ‘threat’ conditioned stimulus (CS+) and ‘safe’ conditioned stimulus (CS-) trials in the order as presented in the task, measured in beats per minute (bpm). ΔHR >0 indicates tachycardia relative to baseline; ΔHR <0 indicates bradycardia relative to baseline. The DMD group had a significant drop in HR from the first CS+ trial to the subsequent trial, the unconditioned ΔHR, ΔHR_UC_ (mean ΔHR_UC_ -6.1 bpm; *P*=.006) but not the Control group (mean ΔHR_UC_ -0.8 bpm; *P*=.99). (**D**) Box plot of ΔHR_UC_, showing a deceleration in HR in the DMD group but not Control group, with a significant difference between groups in univariate analysis of variance: mean difference 5.6 bpm (95% CI 0.51,16.9; *P*=.04). Error bars in line graphs indicate 95% Confidence Intervals (DMD solid, Control dashed). Solid bars in box plots indicate median values; error bars show the interquartile range calculated with inclusive median; outliers indicated with markers.

The unconditioned change in HR after the initial threat stimulus, ΔHR_UC_, showed a significant deceleration in heart rate in the DMD group, with a mean fall in HR of -6.1bpm (95% CI - 10.7,-1.6; *P*=.006), whilst in the Control group there was no significant change (mean ΔHR_UC_-0.8bpm (95% CI -7.1,5.8), *P*=.99), indicating a significant bradycardic response to threat in the DMD group only (Fig. 3). There was a significant difference of 5.6bpm in mean ΔHR_UC_ between the DMD and Control groups (95% CI 0.51,16.9; *P*=.04; Table 3).

When unconditioned physiological responses, SCR_UC_ and ΔHR_UC_, were compared with parent-report neuropsychiatric symptom scores (anxiety, internalising problems, externalising problems, social communication disorders, inattention, hyperactivity), there were significant positive correlations of Anxiety and Internalising problems (incorporating anxiety and depression) scores with SCR_UC_ (Anxiety vs. SCR_UC_: *ρ*=0.38, *P*=.01; Internalising vs. SCR_UC_: *ρ*=0.33, *P*=.03; Supplemental Table 2). In contrast, the measures relating to neurodevelopmental problems (inattention, hyperactivity and social communication problems) did not correlate with SCR_UC_, suggesting that this association is specific to anxiety alone and not related to more general psychopathology. This supports the validity of using the unconditioned skin conductance response as a biological correlate for trait anxiety. ΔHR_UC_ did not significantly correlate with any neuropsychiatric outcomes.

#### Similar acquisition of conditioned responses in both groups

Both DMD and Control groups showed significant discrimination between CS+ and CS-for all Acquisition blocks, for example for Acquisition block 1, DMD mean difference in SCR_CS+_ and SCR_CS-_ was 2.80µS (95% CI 2.6,3.0; *P*<.001) and Control mean difference 1.7µS (95% CI 1.5,2.0; *P*<.001) (Supplemental Table 3). This indicates that participants in both groups successfully discriminated between ‘threat’ and ‘safe’ cues and learned the association with the ‘threat’ stimulus, CS+.

The degree of acquisition of the conditioned response was determined from the differential SCR, SCR_Diff_ (the difference between SCR_CS+_ and SCR_CS-_ in contiguous trials), in the First Interval Response (FIR) window (Fig. 5A). This window captured responses occurring 0-6s after CS presentation, before the ‘threat’ stimulus was presented, thus allowing responses to the CS alone to be identified. There was no difference in SCR_Diff_ between groups in the FIR window, suggesting that fear learning was acquired similarly in both groups (ACQ1 Mean difference -0.14 (−0.71,0.42), *P*=.62; ACQ2 Mean difference 0.17 (−0.29,0.63), *P*=.45; ACQ3 Mean difference -0.21 (−0.66,0.24), *P*=.35).

**Figure 5.**
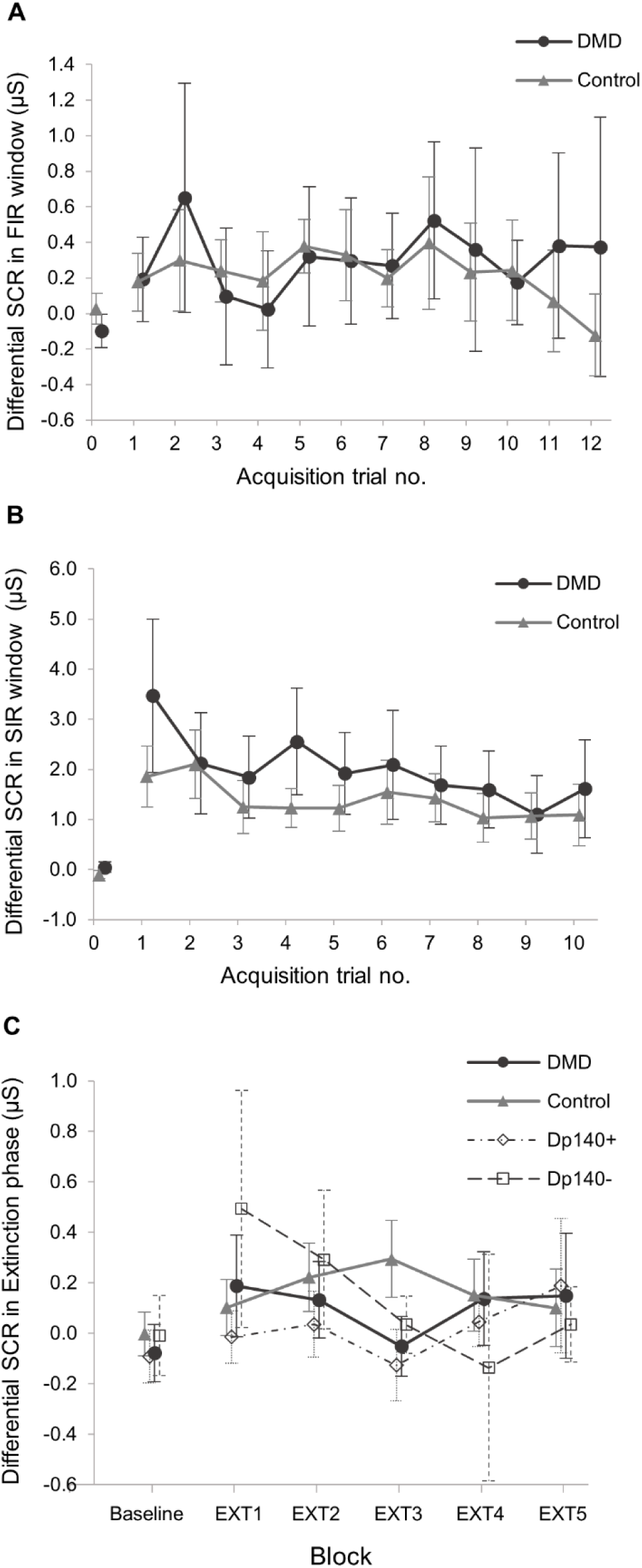
Conditioned response acquisition, habitutation to unconditioned stimulus and retention of conditioned responses. Skin conductance responses (SCR) shown as the differential SCR (SCR_Diff_) to represent the degree of discrimination between ‘threat’ conditioned stimulus (CS+) and ‘safe’ conditioned stimulus (CS-) cues. SCR_Diff_ = SCR_CS+_ - SCR_CS-_ in contiguous trial pairs; SCR_Diff_ = 0 indicates no difference in response to the ‘threat’ CS+ and ‘safe’ CS- cues. **(A)** Mean SCR_Diff_ in the First Interval Response (FIR) window (0-6 seconds after CS onset) for DMD and Control groups. This shows SCR_Diff_ after CS presentation but before the aversive noise, indicating the degree of learned response acquisition. For acquisition trial no. 1, the FIR measurement occurred before participants had been presented with the first aversive stimulus, therefore this is lower than for the subsequent trials in both groups. (**B**) Mean SCR_Diff_ in the Second Interval Response (SIR) window (6-12 seconds after CS onset, the start of which corresponds to the onset of the aversive ‘threat’ stimulus) for DMD and Control groups. These are unconditioned responses to the aversive stimulus, which are shown to reduce or ‘habituate’ with repeated presentations of the aversive stimulus. For SIR data, one trial pair in each of Acquisition blocks 2 and 3 was omitted as these included unreinforced CS+ trials (no aversive noise presented). (**C**) Mean SCR_Diff_ in the Extinction phase blocks for Control and DMD groups, and DMD isoform subgroups (Dp140+ and Dp140-). Extinction of the conditioned response (SCR_CS+_) is defined at the point at which there is no discrimination between the ‘threat’ CS+ and ‘safe’ CS+ cues, i.e. when SCR_Diff_ = 0. Extinction data were extracted from the whole response window (0-12s after CS+ presentation). Mean SCR for familiarisation phase is included as a baseline comparison in all plots. Error bars show 95% Confidence Intervals for each data point.

The Second Interval Response window (SIR; 6-12s after CS onset; Fig. 5B) was timed to capture the response to the unconditioned ‘threat’ stimulus. SCR_Diff_ in the SIR decreased with repeated presentation of the ‘threat’ stimulus in both groups, a phenomenon known as habituation. This could also be seen by comparing SCR_CS+_ between Acquisition blocks 1 and 3 within each group, which showed a significant reduction in both groups (DMD mean difference -1.2µS (−1.7,-0.80), *P*<.001; Control mean difference -0.65µS (−1.1,-0.23), *P*<.001). Habituation is a well-described phenomenon and indicates a reduced salience of the threat stimulus with repeated presentations.^49-51^

#### Retention and extinction of conditioned skin conductance responses

Neither DMD nor Control groups exhibited retention of the conditioned response in the first Extinction block, with no evident discrimination between SCR_CS+_ and SCR_CS-_ in either group (DMD mean difference 0.18µS (95% CI -0.1,0.4), *P*=.21; Control mean difference 0.09µS (95% CI -0.2,0.4), *P*=.52) and no difference in SCR_EXT_ in the first Extinction trial between DMD and Control groups (mean difference 0.37µS (95% CI -0.23,0.96), *P*=.22)(Table 3).

There was a trend of extinction in the DMD group by the third Extinction block, potentially indicating successful conditioned response extinction, which was not evident in the Control group (Fig. 5C). However, this trend was apparent only in the Dp140-isoform subgroup, which showed retention of the conditioned response with a significantly higher SCR_EXT_ compared to Controls (mean difference 1.1µS (95% CI 0.2,2.1), *P*=.02), but not between the Dp140+ subgroup and Controls (mean difference 0.34µS (95% CI -0.2,0.9), *P*=.22) (Table 3). The difference in SCR_EXT_ between the Dp140+ and Dp140-subgroups was non-significant (mean difference 0.84µS (95% CI -0.1,1.8), *P*=.08), although this study was not powered to investigate isoform subgroup differences.

Overall, there were no differences between DMD and Controls in retention or extinction of the conditioned startle response, however the subgroup results suggest that there may be an isoform effect of Dp140 on conditioning.

## Discussion

In recent years there has been an increasing understanding of the role that the multiple dystrophin isoforms have in brain function, both in human and various animal models.^10,11,52-54^ However, some aspects of the complex neurobehavioural phenotype are only now beginning to be better understood. There is growing evidence to suggest that dystrophin is implicated in fear and stress responses in experimental and naturally occurring animal models, including the exaggerated startle response in the *mdx* mouse,^18^ but until now there has been no systematic study of equivalent responses in humans with DMD. This study is the first to obtain objective evidence for a pathological unconditioned startle response in boys with DMD using a psychophysiological fear-conditioning task.

We found increased anxiety symptoms in DMD, with higher scores on measures of anxiety and internalising problems in the DMD compared to the Control group. Furthermore, in comparing to population normative data, the DMD subgroup lacking the Dp140 dystrophin isoform (Dp140-) had significantly higher anxiety scores than the normal population, whilst the subgroup retaining the Dp140 isoform (Dp140+) did not. Previous studies have also shown higher anxiety prevalence for all genotypes of DMD than the typical population, and whilst some observed that anxiety was more prevalent in those with more distal 3’ mutations, in these studies the genotype stratifications did not allow for precise definition of whether the Dp140 isoform was expressed or not.^10-12^

Anxiety is the anticipation of a perceived future threat, associated with more long-lasting increased arousal and apprehension,^55^ whilst fear is a rapid-onset emotional response to an immediate threat, mediated by the amygdala, and is well-conserved across vertebrate species.^56,57^ Anxiety disorders in humans are thought to be caused by excessive activation in innate fear circuits.^29^ In fear-conditioning tasks anxious individuals typically show increased unconditioned startle responses compared to non-anxious individuals,^44,45^ and greater retention of conditioned responses in extinction.^58^

The pattern of psychophysiological responses in the DMD group in our study has similarities with that seen in anxiety disorders, as the DMD group had significantly greater startle responses than Controls, most notably for the unconditioned skin conductance response (SCR_UC_) but also the change in heart rate response (ΔHR_UC_). SCR_UC_, but not ΔHR_UC_, also correlated with anxiety symptom scores (anxiety and internalising problems) in the whole cohort, suggesting SCR_UC_ is a valid physiological correlate of trait anxiety. The increased startle responses we have demonstrated in this study occurred for all genotypes of DMD, irrespective of whether the *DMD* mutation also affects the expression of the shorter Dp140 isoform, and therefore can be attributed solely to the loss of the full-length Dp427 isoform. This is consistent with animal studies: the *mdx* mouse, lacking only Dp427 dystrophin, displays an abnormal unconditioned startle or ‘fear’ response. We therefore conclude that the SCR_UC_ startle response is a useful biomarker of a deficiency of the full-length Dp427 dystrophin isoform.

Whilst there was no apparent effect of Dp140 isoform presence or absence on the startle response magnitude, the anxiety score profile was different in the Dp140 isoform subgroups when compared to the normal population, with higher anxiety in the Dp140-subgroup. Increased retention of the conditioned SCR compared to Controls also only occurred in the Dp140- and not the Dp140+ subgroup, which is a typical feature of anxiety disorders.^58,59^ Increased response retention in Dp140-may also relate to increased salience of auditory stimuli, given the increased tendency to neurodevelopmental symptoms in this group that can be associated with sensory over-reactivity.^60^ Furthermore, half of those lacking the Dp140-isoform terminated the Extinction phase early, which could be attributed to heightened arousal. Given the small isoform subgroup sizes we cannot draw definite conclusions from these findings, and to date there are no reported behavioural studies of startle responses in mice lacking the Dp140 isoform to use as a comparison. However, our data indicate that while the deficiency of Dp427, a feature of all DMD boys, is associated with increased anxiety prevalence and heightened activity in fear circuits, mutations located towards the 3’ end that also affect the Dp140 isoform may further exacerbate these symptoms. This warrants attention in future studies.

GABAergic synapse dysfunction has been demonstrated in the *mdx* mouse, lacking full length Dp427 dystrophin. In wild-type mice, GABA_A_-receptors (GABA_A_-R) on post-synaptic membranes co-localise with dystrophin at inhibitory synapses in the hippocampus, cerebellum, and amygdala.^61^ In the *mdx* mouse there is reduced clustering of GABA_A_-Rs and disrupted synaptic function in the basolateral amygdala and hippocampus.^19,61,62^ CNS dystrophin-restoration (either by direct injection or viral vector administration) of antisense oligonucleotides (AON)^18,19,24,63^ corrects both the synaptic abnormalities and the exaggerated startle response. In a different mouse model with a heterozygous mutation in the γ_2_-GABA_A_-R subunit, reduced GABA_A_-receptor clustering also occurs and is associated with heightened responses in fear-conditioning tasks, increased harm avoidance behaviour and an explicit memory bias to threat cues.^64^ These findings suggest that reduced post-synaptic GABA_A_-R density is a key factor in the enhanced fear phenotype.

In typical humans, dystrophin is expressed in the so-called ‘limbic’ structures, including the amygdala, the hippocampal formation and the parahippocampal gyrus.^3,56,65^ Imaging studies have suggested limbic dysfunction occurs in DMD: hippocampal and medial temporal lobe hypometabolism has been observed by FDG-PET^66^, hippocampal hyperconnectivity is seen in default mode network fMRI^67^ and GABA_A_-receptor distribution is abnormal in the prefrontal cortex.^20^ Dysfunction at GABAergic inhibitory synapses caused by loss of Dp427 dystrophin may therefore underlie the pathogenesis of at least some of the complex neuropsychiatric phenotype in DMD. Impaired GABAergic transmission has been implicated in the pathogenesis of autism spectrum disorder and depression.^68,69^ Accordingly, it is possible that some mental health issues associated with DMD are potentially reversible in humans following CNS-targeted dystrophin restoration, as in the *mdx* mouse.

The outlook for individuals with DMD now is very different to that in previous decades. Following improved standards of care, life-expectancy has increased by approximately 10 years,^70,71^ and genetic therapies addressing dystrophin deficiency in muscle are likely to further improve functional status and survival. However, the neuropsychiatric aspects of DMD impact on the daily functioning of individuals living with DMD and increase carer-burden.^72-75^ Current care recommendations advise consideration of neuropsychological referral at diagnosis of DMD, and if neurodevelopmental problems arise,^72^ although there is no specific guidance for assessment, monitoring or treatment of psychiatric disorders and specific treatment for psychiatric disorders in DMD is uncommon.^74^ Therefore, it is increasingly important to identify and manage neuropsychiatric symptoms with therapeutic interventions, and potentially with future disease-modifying CNS-acting therapies.

CNS-targeted intrathecal AON therapy is effective in spinal muscular atrophy,^76^ however in DMD systemic AON therapy is the primary requirement to ameliorate the muscle pathology. The current peripherally-administered AONs in DMD do not cross the blood-brain barrier, but systemically-delivered AONs with improved CNS penetration are under development,^24,77^ which could potentially enable dystrophin restoration in muscle and CNS from the same systemically-administered therapy.

We acknowledge several limitations in this study. As this was the first study of its kind in this clinical population, sample sizes were based on previous literature and pragmatic considerations as ‘a priori’ power calculations were not possible. However, data from this study will be used to inform the design of future studies. Isoform subgroups analyses were exploratory outcomes and subgroup sizes were underpowered, therefore the subgroup findings should be interpreted with caution, however as DMD is a rare disorder and some genotypes are uncommon it can be practically difficult to recruit to all genotype subgroups. We compared DMD participants against age- and sex-matched healthy control subjects in this proof-of-principle investigation, however we did not control for steroid use or physical disability, which would have eliminated further potentially confounding factors. Artefacts and early termination led to data loss, most notably in the Extinction phase, which limited data interpretation for this phase. Retention of conditioned responses may have been improved if the extinction task was delayed, as retention has been reported to be optimal after 24 hours,^78^ however for practical reasons all task phases were performed on the same day to limit the number of visits, which could be more difficult especially for DMD participants. Selection bias may have occurred in the Control group, given the higher FSIQ and anxiety scores compared to the normal population, and in the DMD group a potential bias towards participants without significant cognitive or neurodevelopmental problems. However, given these potential biases, it is reassuring that between-group differences in the primary outcomes were still identified. The emotional and behavioural screening instruments used are not diagnostic measures and may have limitations in DMD,^79^ therefore caution should be taken in interpreting neuropsychiatric data.

In conclusion, we have demonstrated for the first time that boys with DMD show increased physiological startle responses to threat, using the principle of ‘back-translation’ from the exaggerated startle response phenotype in *mdx* mice. This has implications both for the further understanding of the neurobiology of DMD and clinical management. We propose that a lack of full-length Dp427 dystrophin leads to a dysfunctional fear system which predisposes individuals with DMD to develop anxiety disorders, and the lack of the Dp140 dystrophin isoform may further increase this risk. To date, anxiety in DMD has received little attention in both research and clinical practice therefore robust prevalence data are lacking, however given the increased risk of anxiety due to underlying neurobiological changes, we suggest that young boys with DMD should be actively monitored for anxiety and emotional problems from an early age to enable support, interventions and treatment where appropriate. Further investigation of anxiety and other neuropsychiatric disorders is needed to inform clinical practice and, where necessary, refine clinical guidelines to optimise the care of young people with DMD. Our findings also have implications for translational research, as to date no CNS biomarker of dystrophin has been identified that could be used in a clinical trial. We propose that the unconditioned skin conductance startle response is a potential physiological biomarker to evaluate CNS dystrophin restoration, analogous to the *mdx* startle response as an outcome in preclinical studies of CNS-targeted antisense oligonucleotide therapies. The findings from this study may therefore inform current clinical management of anxiety disorders in DMD, and potentially be a first step towards evaluating future CNS-disease modifying therapies.

## Supporting information

Reporting guidelines_STROBE Checklist

## Data Availability

The authors confirm that the data supporting the findings of this study are available within the article and its Supplementary material. In view of the rarity of DMD and that genotype data is included, only aggregated data will be made available on reasonable request.

## Abbreviations

ADHD: Attention Deficit Hyperactivity Disorder
AON: Antisense oligonucleotide
ASD: Autism Spectrum Disorder
Bpm: Beats per minute
CS: Conditioned stimulus
DMD: Duchenne muscular dystrophy
EDA: Electrodermal activity
FIR: First Interval Response
FSIQ: Full Scale Intelligence Quotient
GOSH: Great Ormond Street Hospital for Children
HR: Heart rate
ICH: UCL Great Ormond Street Institute of Child Health
ITI: Inter-trial interval
SCR: Skin conductance response
SIR: Second Interval Response
SPECT: Single positron emission computed tomography
UCL: University College London
UCS: Unconditioned stimulus
UTR: Untranslated region.

## Acknowledgements

We would like to acknowledge the support of the National Institute of Health Research (NIHR) Biomedical Research Centre at Great Ormond Street Hospital for Children NHS Foundation Trust. The views expressed are participants of the author(s) and not necessarily participants of the NHS, the NIHR or the Department of Health. We gratefully acknowledge the GOSH Somers Clinical Research Facility, the participation of Muscular Dystrophy UK, Duchenne UK and Action Duchenne in promoting study recruitment and all the study participants and their parents/carers. Advice on developing the fear conditioning paradigm was provided by David Watson (University of Sussex), Charles Marshall (UCL) & Rachael Elward (UCL).

