## Supplemental Fig. 1 for "Startle responses in Duchenne muscular dystrophy: a novel biomarker of brain dystrophin deficiency"

**Supplemental Figure 1. Flow Diagram of study recruitment.**

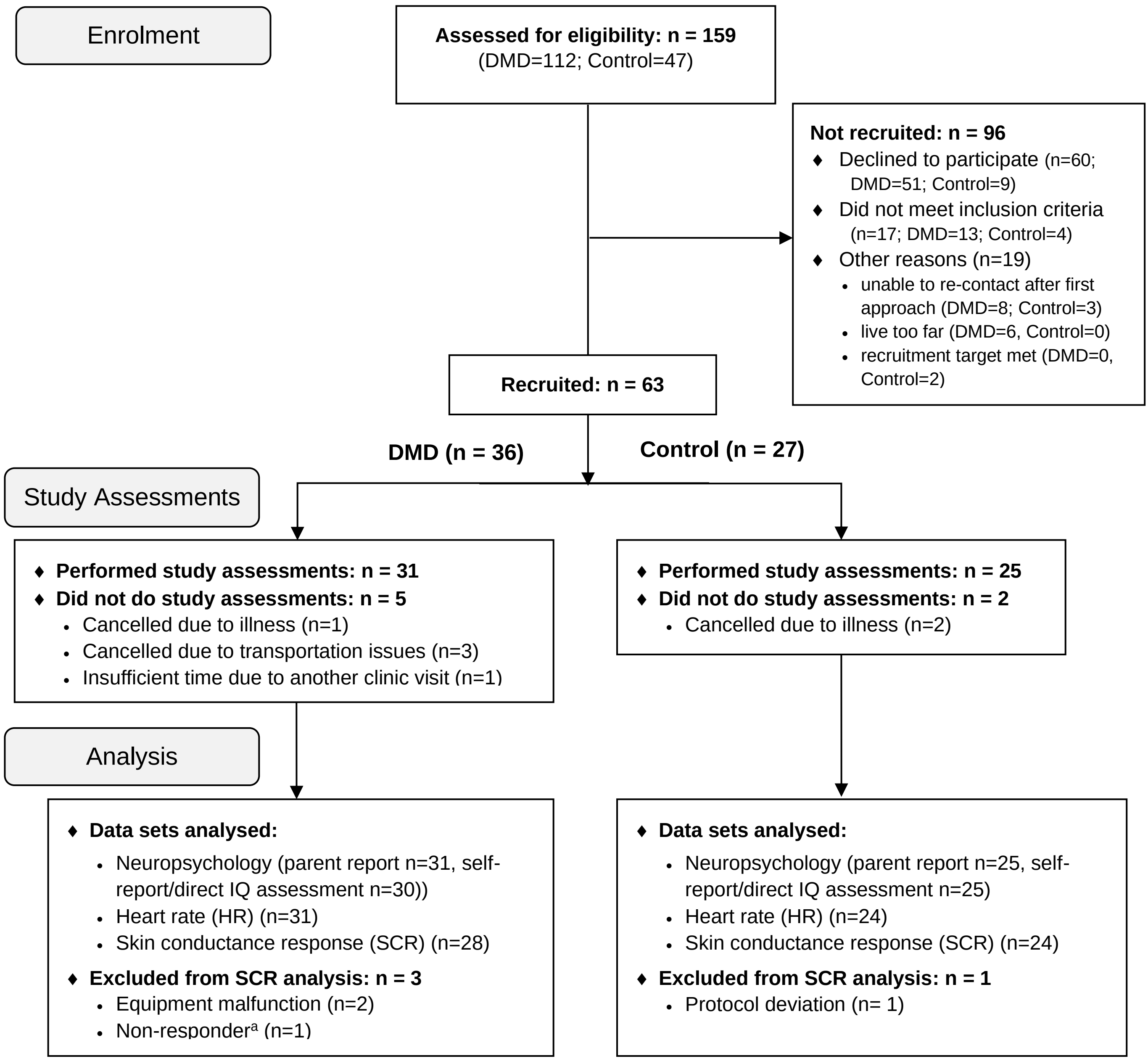


[a]Non-responders in skin conductance recordings were defined as participants in whom ≥ 50% raw SCRs following UCS presentations and pre-exposure tasks were <0.01 µS, based on recommendations from previous literature (Lonsdorf et al, 2019; Marin et al 2019; Hu et al 2019).
