## Supplemental Fig. 2 for "Startle responses in Duchenne muscular dystrophy: a novel biomarker of brain dystrophin deficiency"

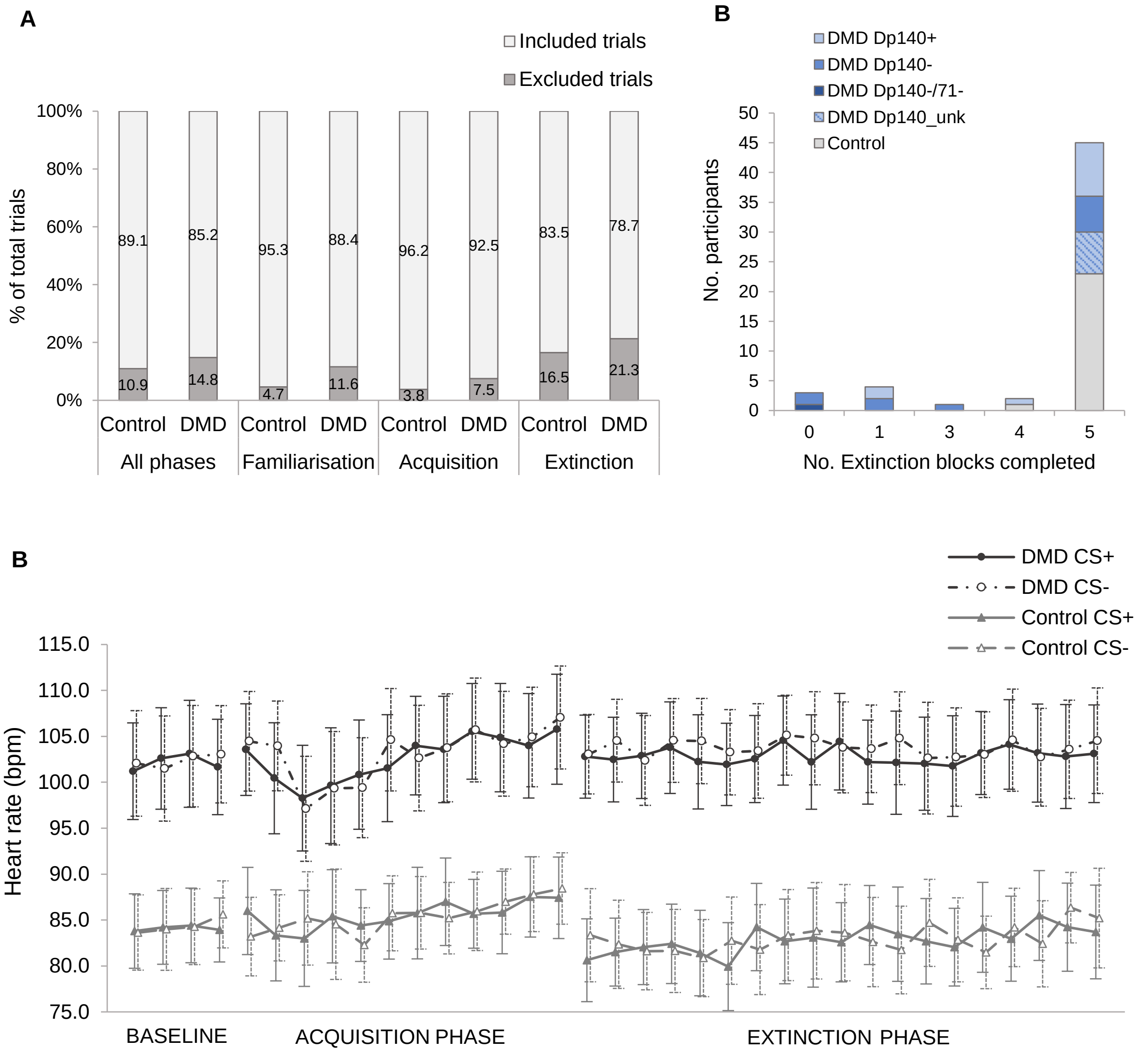


**Supplemental Figure 2. Data exclusion and drop-out.** (**A**) The percentage of individual SCR trial data excluded from analysis due to artefacts. (**B**) No. of Extinction blocks (8 trials each) completed by DMD (n=28) and Control (n=24) participants included in SCR analysis. DMD group is split into isoform subgroups. 24/24 (100%) of Control and 20/28 (71.4%) DMD participants completed 4 or more extinction blocks. Of the 8 DMD ‘Non-completers’ (completing <4 blocks, 6/8 (75%) lacked the Dp140 isoform (5 Dp140-; 1 Dp140-/71-) and 2/8 retained it (Dp140+).
