## Supplemental Fig. 3 for "Startle responses in Duchenne muscular dystrophy: a novel biomarker of brain dystrophin deficiency"

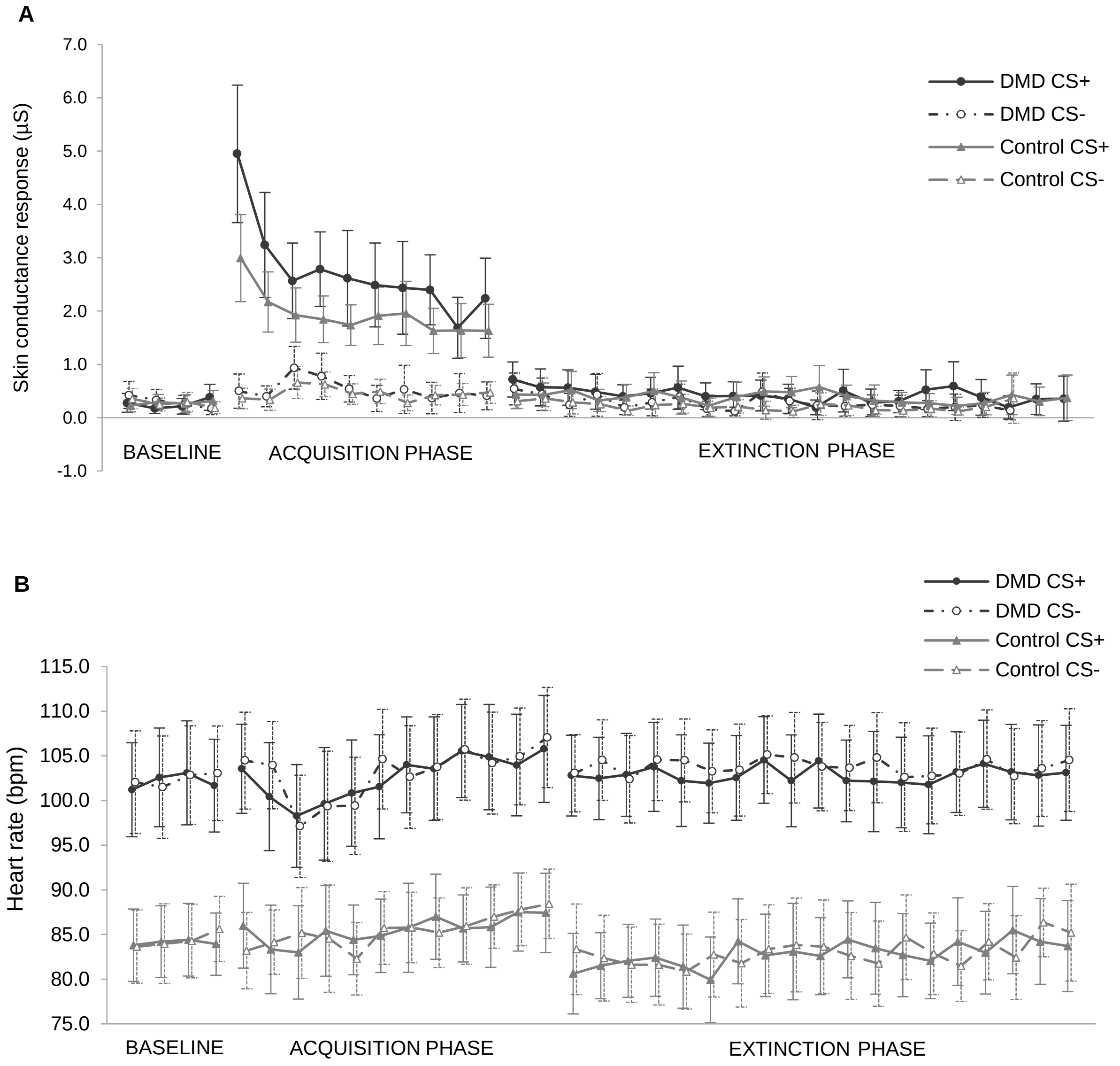


**Supplemental Figure 3: Skin conductance response and heart rate data for each CS+ and CS- trial in all phases of the task.** (**A**) Skin conductance responses (SCR, measured in microSiemens, µS) for DMD and Control groups in CS+ and CS- trials. (**B**) Absolute heart rate (HR, measured in beats per minute, bpm). Baseline is the initial familiarisation phase where both CS+ and CS- are presented for four trials each but without the aversive unconditioned stimulus. Acquisition phase comprises 24 trials (12 of each CS+ and CS-), and Extinction phase comprises 40 trials. Error bars in show 95% Confidence Intervals for each data point.
