## Supplemental Table 1 for "Startle responses in Duchenne muscular dystrophy: a novel biomarker of brain dystrophin deficiency"

**Supplemental Table 1. Neuropsychological assessment mean scores and group comparisons**

|  |  | **Between group comparisons**^b^ | | | | | **Group vs. normative data comparisons**^c^ | | | |
| --- | --- | --- | --- | --- | --- | --- | --- | --- | --- | --- |
| **Neuropsychological assessment**^a^ | **Group** | **n** | **Mean raw score *(SD)*** | **Mean difference *(95% CI)*** | ***t*** | **Sig., *P*** | **Normative data mean score *(SD)*** | **Mean difference**  ***(95% CI)*** | ***t*** | **Sig., *P*** |
| **Full Scale Intelligence Quotient (FSIQ)** | **Control** | 25 | 115.4 *(14.4)* | -24.9  *(-32.5,-17.4)* | -6.7 | **<.001** | 101.4  *(13.0)* | 14.0  *(8.0, 19.9)* | 4.9 | **<.001** |
|  | **DMD** | 30 | 90.5 *(13.2)* |  |  |  |  | -10.9  *(-15.8,-6.0)* | -4.5 | **<.001** |
|  | **DMD_Dp140+** | 12 | 96.4 *(14.1)* | -13.6  *(-25.3,-1.9)* | -2.41 | **.03** |  | -5.0  *(-14.0,4.0)* | -1.2 | .25 |
|  | **DMD_Dp140-** | 11 | 82.8 *(12.8)* |  |  |  |  | -18.6  *(-27.2,-10.0)* | -4.8 | **<.001** |
| **Verbal Comprehension Index (VCI)** | **Control** | 25 | 113.5 *(13.2)* | -21.4  *(-28.4,-14.3)* | -6.1 | **<.001** | 101.2  *(12.7)* | 12.3  *(6.9, 17.7)* | 4.7 | **<.001** |
|  | **DMD** | 30 | 92.2 *(12.6)* |  |  |  |  | -9.0  *(-13.8,-4.3)* | -3.9 | **<.001** |
|  | **DMD_Dp140+** | 12 | 97.3 *(12.9)* | -13.3  *(-23.9,-2.8)* | -2.6 | **.02** |  | -3.9  *(-12.1, 4.3)* | -1.0 | .32 |
|  | **DMD_Dp140-** | 11 | 84.0 *(11.4)* |  |  |  |  | -17.2  *(-24.9,-9.5)* | -5.0 | **<.001** |
| **Performance Reasoning Index (PRI)** | **Control** | 25 | 113.8 *(15.8)* | -23.1  *(-4.2,-31.4)* | -5.5 | **<.001** | 101.2  *(12.5)* | 12.6  *(6.1, 19.2)* | 4.0 | **<.001** |
|  | **DMD** | 30 | 90.8 *(15.0)* |  |  |  |  | -10.4  *(-16.0,-4.8)* | -3.8 | **<.001** |
|  | **DMD_Dp140+** | 12 | 95.9 *(16.9)* | -10.8  *(-24.8,3.1)* | -1.6 | .12 |  | -5.3  *(-16.0, 5.4)* | -1.1 | .30 |
|  | **DMD_Dp140-** | 11 | 85.1 *(15.2)* |  |  |  |  | -16.1  *(-26.3,-5.9)* | -3.5 | **<.001** |
| **Anxiety score (parent-report)** | **Control** | 25 | 12.0 *(8.1)* | 5.4  *(0.09, 10.8)* | 2.0 | **.046** | 12.1  *(12.4)* | -0.06  *(-3.4, 3.3)* | -0.04 | .97 |
|  | **DMD** | 31 | 17.5 *(11.2)* |  |  |  |  | 5.4  *(1.3, 9.5)* | 2.7 | **.01** |
|  | **DMD_Dp140+** | 12 | 16.7 *(11.3)* | 3.1  *(-5.4, 11.5)* | 0.75 | .46 |  | 4.6  *(-2.6, 11.7)* | 1.4 | .19 |
|  | **DMD_Dp140-** | 11 | 19.7 *(7.7)* |  |  |  |  | 7.6 *(2.5, 12.8)* | 3.3 | **.008** |
| **Anxiety score (self-report)** | **Control** | 25 | 19.9 *(9.2)* | -2.9  *(-7.7, 1.9)* | -1.2 | .22 | 12.7  *(9.4)* | 7.2  *(3.4, 11.0)* | 3.9 | **<.001** |
|  | **DMD** | 30 | 16.9 *(8.5)* |  |  |  |  | 4.2  *(1.1, 7.4)* | 2.7 | **.01** |
|  | **DMD_Dp140+** | 12 | 18.4 *(7.9)* | -2.3  *(-9.1, 4.6)* | -0.7 | .49 |  | *5.7*  *(0.7, 10.8)* | 2.5 | **.03** |
|  | **DMD_Dp140-** | 11 | 16.1 *(8.0)* |  |  |  |  | 3.4  *(-2.0, 8.8)* | 1.4 | .19 |
| **Internalising problems** | **Control** | 25 | 6.5 *(4.7)* | 4.6  *(1.2, 7.9)* | 2.7 | **.009** | 5.1  *(4.8)* | 1.4  *(-0.6, 3.3)* | 1.5 | .16 |
|  | **DMD** | 31 | 11.0 *(7.3)* |  |  |  |  | 5.9  *(3.3, 8.6)* | 4.6 | **<.001** |
|  | **DMD_Dp140+** | 12 | 8.8 *(4.5)* | 4.3  *(-1.5, 10.0)* | 1.6 | .14 |  | 3.7  *(0.8, 6.5)* | 2.8 | **.02** |
|  | **DMD_Dp140-** | 11 | 13.0 *(7.9)* |  |  |  |  | 7.9  *(2.6,13.2)* | 3.3 | **.008** |
| **Externalising problems** | **Control** | 25 | 5.5 *(7.1)* | 6.5  *(0.9, 12.2)* | 2.3 | **.02** | 6.6  *(6.0)* | -1.1  *(-4.0, 1.8)* | -0.8 | .45 |
|  | **DMD** | 31 | 12.1 *(12.4)* |  |  |  |  | 5.5  *(0.9, 10.0)* | 2.4 | **.02** |
|  | **DMD_Dp140+** | 12 | 9.9 *(8.6)* | 4.3  *(-7.7, 16.3)* | 0.7 | .47 |  | 3.3  *(-2.2, 8.8)* | 1.3 | .21 |
|  | **DMD_Dp140-** | 11 | 14.2 *(17.9)* |  |  |  |  | 7.6  *(-4.4, 19.6)* | 1.4 | 0.19 |
| **Social communication problems** | **Control** | 25 | 3.6 *(4.6)* | 4.8  *(1.9, 7.8)* | 3.2 | **.003** | 3.3  *(4.2)* | 0.3  (-1.6, 2.2) | 0.3 | .75 |
|  | **DMD** | 30 | 8.4 *(6.4)* |  |  |  |  | 5.1  *(2.8, 7.5)* | 4.4 | **<.001** |
|  | **DMD_Dp140+** | 12 | 6.2 *(5.2)* | 4.4  *(-0.9, 9.6)* | 1.7 | .10 |  | 2.9  *(-0.5, 6.2)* | 1.9 | .08 |
|  | **DMD_Dp140-** | 11 | 10.6 *(6.9)* |  |  |  |  | 7.2  *(2.6, 11.9)* | 3.5 | **.006** |
| **Inattention** | **Control** | 25 | 4.1 *(3.2)* | 2.0  *(0.06, 4.0)* | 2.1 | **.04** | 3.0  *(2.8)* | 1.1  *(-1.9, 2.4)* | 1.8 | .09 |
|  | **DMD** | 31 | 6.2 *(4.0)* |  |  |  |  | 3.2  *(1.7, 4.6)* | 4.4 | **<.001** |
|  | **DMD_Dp140+** | 12 | 4.1 *(3.8)* | 3.9  *(0.6, 7.2)* | 2.5 | **.02** |  | 1.1  *(-1.3, 3.5)* | 1.0 | .34 |
|  | **DMD_Dp140-** | 11 | 8.0 *(3.9)* |  |  |  |  | 5.0  *(2.4, 7.6)* | 4.3 | **.002** |
| **Hyperactivity** | **Control** | 25 | 4.3 *(4.3)* | -0.12  *(-2.3, 2.1)* | -0.1 | .91 | 2.5  *(2.9)* | 1.8  *(0.02, 3.5)* | 2.1 | **0.047** |
|  | **DMD** | 31 | 4.2 *(3.9)* |  |  |  |  | 1.7  *(0.2, 3.1)* | 2.4 | **0.02** |
|  | **DMD_Dp140+** | 12 | 2.7 *(2.8)* | 3.4  *(0.2, 6.7)* | 2.2 | **.04** |  | 0.2  *(-1.6, 2.0)* | 0.2 | 0.84 |
|  | **DMD_Dp140-** | 11 | 6.1 *(4.6)* |  |  |  |  | 3.6  *(0.5, 6.7)* | 2.6 | **0.03** |

^a^ Neuropsychological/neuropsychiatric assessments using either direct assessment (FSIQ/VCI/PRI), parent-report or self-report (Anxiety only) questionnaires: FSIQ-4/VCI/PRI - Wechsler Abbreviated Intelligence Scale – 2nd Edition; Anxiety - Screen for Child Anxiety Related Disorders Parent-report/Child-report; Internalising and Externalising problems - Child Behavior Checklist (CBCL); Social communication problems - Social Communication Disorders Checklist; Inattention and Hyperactivity - Conners’ Parent Rating Scale-Revised, short version.

^b^ Between group comparisons performed with independent samples *t*-tests, showing mean difference, standard deviation (SD) and 95% confidence interval of the difference (95% CI); *t* test statistic. Sig. = Two-tailed significance, using alpha level of *P*=.05. *P*-values <.05 are highlighted in bold

^c^ Group data compared to published age-matched population normative data for each instrument using one-sample *t*-tests. FSIQ-4/VCI/PRI normative data from WASI-II manual (Wechsler, D. (1999). Wechsler Abbreviated Scale of Intelligence. San Antonio, TX: The Psychological Corporation); Internalising/Externalising normative data from CBCL manual (Achenbach & Rescorla, 2001); Screen for Child Anxiety Related Disorders (SCARED) normative data from a large US population study (Sequeira et al, 2019); Social Communication Disorders Checklist normative data from a large UK population study (Skuse et al 2009); Conners’ Parent Rating Scale-Revised normative data from published manual (Conners CK, Sitarenios G, Parker JDA, Epstein JN. The revised Conners' Parent Rating Scale (CPRS‐R): factor structure, reliability, and criterion validity. Journal of Abnormal Child Psychology 1998;26(4):257‐68). Normative data was available for: mean raw score, standard deviation (except SCARED) and sample size; males only (except SCARED which includes males and females); and was age-matched, except SCARED (11-12 years only) and SCDC (7-8 years only).
