## Supplemental Table 2 for "Startle responses in Duchenne muscular dystrophy: a novel biomarker of brain dystrophin deficiency"

**Supplemental Table 2. Correlation of neuropsychiatric measures with (A) full-scale IQ and (B) primary unconditioned physiological responses.**

| **A Correlation coefficients for full-scale IQ vs. neuropsychiatric and physiological outcomes** | | | | | | | | | | | | | | | | | |
| --- | --- | --- | --- | --- | --- | --- | --- | --- | --- | --- | --- | --- | --- | --- | --- | --- | --- |
|  | | **Anxiety** | | **Internalising** | | **Externalising** | | **Social communication** | | **Inattention** | | **Hyperactivity** | | **SCR_UC_**^c^ | | **ΔHR_UC_**^d^ | |
|  | | ***rho***^a^ | ***P***^b^ | ***rho*** | ***P*** | ***rho*** | ***P*** | ***rho*** | ***P*** | ***rho*** | ***P*** | ***rho*** | ***P*** | ***rho*** | ***P*** | ***rho*** | ***P*** |
|  | All (n=56)  DMD (n=31)  Control (n=25) | **-0.30** | ***0.03*** | **-0.30** | ***0.03*** | **-0.30** | ***0.03*** | **-0.35** | ***0.009*** | **-0.38** | ***0.004*** | 0.069 | *0.62* | -0.003 | *0.98* | -0.02 | *0.87* |
|  |  | -0.04 | *0.83* | 0.07 | *0.71* | -0.01 | *0.98* | -0.14 | *0.46* | **-0.42** | ***0.02*** | -0.05 | *0.78* | 0.21 | *0.29* | -0.16 | *0 .42* |
|  |  | -0.18 | *0.4* | -0.15 | *0.49* | 0.02 | *0.93* | 0.14 | *0.52* | -0.03 | *0.89* | 0.39 | *0.05* | 0.386 | *0.06* | -0.31 | *0.14* |

| **B Correlation coefficients for unconditioned physiological responses vs. neuropsychiatric symptom scores** | | | | | | | | | | | | | |
| --- | --- | --- | --- | --- | --- | --- | --- | --- | --- | --- | --- | --- | --- |
|  |  | **Anxiety** | | **Internalising** | | **Externalising** | | **Social communication** | | **Inattention** | | **Hyperactivity** | |
| **i. Adjusted for IQ** | | ***ρ***^e^ | ***P*** | ***ρ*** | ***P*** | ***ρ*** | ***P*** | ***ρ*** | ***P*** | ***ρ*** | ***P*** | ***ρ*** | ***P*** |
| SCR_UC_^c^ | All (n=47) | **0.38** | **.01** | **0.33** | **.03** | 0.13 | .40 | 0.27 | .07 | 0.05 | .74 | 0.11 | .45 |
|  | DMD (n=31) | 0.37 | .08 | 0.27 | .22 | 0.17 | .44 | 0.33 | .13 | 0.10 | .65 | 0.19 | .39 |
|  | Control (n=25) | 0.22 | .32 | 0.04 | .86 | -0.07 | .76 | -0.09 | .68 | -0.05 | .83 | 0.01 | .96 |
| ΔHR_UC_^d^ | All (n=56) | 0.02 | .91 | -0.05 | .75 | -0.15 | .32 | -0.13 | .37 | -0.02 | .92 | -0.001 | .99 |
|  | DMD (n=31) | -0.07 | .77 | 0.03 | .88 | -0.20 | .36 | -0.02 | .94 | -0.02 | .92 | 0.002 | .99 |
|  | Control (n=25) | 0.23 | .31 | 0.10 | .65 | -0.07 | .75 | -0.15 | .51 | 0.008 | .97 | 0.01 | .95 |
| **ii. Unadjusted for IQ** | | ***rho*** | ***P*** | ***rho*** | ***P*** | ***rho*** | ***P*** | ***rho*** | ***P*** | ***rho*** | ***P*** | ***rho*** | ***P*** |
| SCR_UC_ | All (n=56) | **0.29*** | **.04** | *0.23* | .10 | *0.18* | .21 | 0.17 | .25 | *-0.10* | .95 | *0.14* | .31 |
|  | DMD (n=31) | 0.28 | .15 | *0.29* | .13 | *0.22* | .26 | 0.21 | .29 | *0.00* | .99 | *0.20* | .31 |
|  | Control (n=25) | 0.12 | .57 | *-0.01* | .96 | *-0.07* | .73 | -0.16 | .45 | *-0.32* | .13 | *-0.02* | .92 |
| ΔHR_UC_ | All (n=56) | 0.10 | .45 | *0.06* | .66 | *-0.21* | .13 | -0.15 | .27 | *-0.12* | .93 | *-0.07* | .60 |
|  | DMD (n=31) | 0.13 | .49 | *0.11* | .56 | *-0.17* | .35 | -0.11 | .56 | *-0.07* | .72 | *-0.14* | .44 |
|  | Control (n=25) | 0.20 | .34 | *0.22* | .30 | *-0.09* | .67 | 0.007 | .97 | *0.24* | .25 | *0.00* | .99 |

^a^ *rho* = Spearman correlation coefficient.

^b^ *P* = signficance, with significance level alpha of *P*=.05. P-values <.05 and corresponding correlation statistics are shown in bold.

^c^ SCR_UC_ = Unconditioned skin conductance response to the first ‘threat’ trial with conditioned stimulus, CS+, in microSiemens (µS).

^d^ ΔHR_UC_ = Unconditioned change in heart rate response to the first CS+ ‘threat’ trial, in beats per minute (bpm).

^e^ *ρ* = Partial correlation coefficient, controlling for full-scale IQ.
