## Supplemental Table 3 for "Startle responses in Duchenne muscular dystrophy: a novel biomarker of brain dystrophin deficiency"

**Supplemental Table 3. Discrimination between CS+ and CS- stimuli determined by mean difference in skin conductance responses (SCR_CS+_ vs. SCR_CS-_).**

| **Mean differences between SCR_CS+_ and SCR_CS-_ in µS by block**^a^ | | | | | | | |
| --- | --- | --- | --- | --- | --- | --- | --- |
|  | | **DMD** | | | **Control** | | |
| **Block** | | **Mean difference**  ***(95% CI)*** | **df** | **Sig., *P***^b^ | **Mean difference *(95% CI)*** | **df** | **Sig., *P*** |
| Familiarisation block | | 0.006 (-0.23, 0.25) | 2812.7 | 0.96 | 0.02 (-0.23, 0.27) | 2810.5 | .86 |
| Acquisition blocks | 1 | **2.80 (2.57, 3.04)** | 2812.7 | **<.001** | **1.7 (1.5, 2.0)** | 2810.3 | **<.001** |
|  | 2 | **2.01 (1.79, 2.33)** | 2810.9 | **<.001** | 1**.4 (1.1, 1.6)** | 2810.5 | **<.001** |
|  | 3 | **1.74 (1.46, 2.01)** | 2811.0 | **<.001** | **1.1 (0.8, 1.4)** | 2811.1 | **<.001** |
| Extinction blocks | 1 | 0.18 (-0.10, 0.43) | 2810.7 | 0.21 | 0.09 (-0.18, 0.35) | 2810.6 | 0.52 |
|  | 2 | 0.16 (-0.12, 0.44) | 2811.2 | 0.27 | 0.14 (-0.14, 0.41) | 2810.3 | 0.33 |
|  | 3 | 0.07 (-0.23, 0.36) | 2811.3 | 0.65 | **0.37 (0.10, 0.64)** | 2811.6 | **0.007** |
|  | 4 | 0.23 (-0.07, 0.54) | 2811.6 | 0.13 | 0.13 (-0.17, 0.42) | 2811.3 | 0.40 |
|  | 5 | 0.21 (-0.11, 0.53) | 2811.2 | 0.20 | 0.15 (-0.14, 0.43) | 2810.7 | 0.32 |

^a^ SCR_CS+_ = skin conductance response in CS+ ‘threat’ trials; SCR_CS-_ = skin conductance response in CS- ‘safe’ trials. The difference between these two values indicates the degree of discrimination between the two trial types, showing the mean difference for each block (eight trials; four of each type) in both DMD and Control groups.

^b^ Statistical analysis conducted using linear mixed models analysis. 95% CI = 95% confidence interval; df = degrees of freedom; Sig. = significance level alpha, *P*=.05. *P*-values <.05 shown in bold.
