## Supplemental Table 4 for "Startle responses in Duchenne muscular dystrophy: a novel biomarker of brain dystrophin deficiency"

| **Primary SCR and HR metric comparisons, not adjusted for IQ** | | | | | | | | | |
| --- | --- | --- | --- | --- | --- | --- | --- | --- | --- |
|  | **DMD vs. Control** | | | **DMD Dp140+ vs. Control** | | | **DMD Dp140- vs. Control** | | |
| **Outcome measure** | **Mean diff. *(95% CI)*** | ***F*** | **Sig., P**^d^ | **Mean diff. *(95% CI)*** | ***F*** | **Sig., *P*** | **Mean diff. *(95% CI)*** | ***F*** | **Sig., *P*** |
| SCR_UC_  (in µS)^a^ | 2.0  *(0.3, 3.6)* | 5.9 | **.02** | 2.4  *(0.4, 4.3)* | 6.2 | **.02** | 2.0  *(0.03, 3.9)* | 4.3 | **.047** |
| ΔHR_UC_  (in bpm)^b^ | -5.6  *(-11.6, 0.4)* | 3.5 | .07 | -6.5  *(-14.8, 1.9)* | 2.5 | .13 | -1.8  *(-10.6, 7.0)* | 0.17 | 0.68 |
| SCR_EXT_  (in µS)^c^ | 0.28  *(-0.16,0.72)* | 1.7 | .20 | 0.17  *(-0.31, 0.65)* | 0.5 | 0.47 | 0.78  *(0.14, 1.4)* | 6.2 | **0.02** |

**Supplemental Table 4. Primary outcome measure between group comparisons, without adjustment for IQ.**

^a^ SCR_UC_ = Unconditioned skin conductance response to the first ‘threat’ trial with conditioned stimulus, CS+, in microSiemens (µS).

^b^ ΔHR_UC_ = Unconditioned change in heart rate response to the first CS+ ‘threat’ trial, in beats per minute (bpm).

^c^ SCR_EXT_ = Conditioned skin conductance response to the first CS+ trial of the Extinction phase, in µS.

^d^ Significance (Sig.) testing using univariate analysis of variance between groups/subgroups, taking alpha of *P*=.05; *P*-values <.05 are highlighted in bold. *F* = effect size of Group/Isoform group. Bonferroni adjustment made for multiple comparisons.
