## Supplemental Table 5 for "Startle responses in Duchenne muscular dystrophy: a novel biomarker of brain dystrophin deficiency"

**Supplemental Table 5. Demographics and baseline neuropsychiatric data for DMD completers and non-completers in extinction phase.**

|  | **Completer**^a^ | **Non-completer**^a^ |  | |
| --- | --- | --- | --- | --- |
| Median no. extinction blocks completed *(range)* | 5 (*4, 5)* | 1 *(0,3)* |  |  |
| Median no. extinction trials completed (range) | 40.0 *(40, 40)* | 9.5 *(0, 24)* |  |  |
| **Participants** | **n *(%)*** | **n *(%)*** |  |  |
| Total DMD participants | 23 (*74.2)* | 8 (*25.8)* |  |  |
| Dp 140+, n | 10 (83.3) | 2 (16.7) |  |  |
| Dp140-, n^a^ | 6 (50.0) | 6 *(50.0)* |  |  |
| Dp140_unk, n | 7 (100.0) | 0 *(0)* |  | |
| **Demographics and baseline neuropsychiatric data** | **Median *(range)*** | **Median *(range)*** | ***U***^d^ | ***Sig., P*** |
| Age, years | 9.84 (*7.4, 12.0)* | 9.3 *(7.1, 11.4)* | 74.0 | .44 |
| Full scale IQ^b^ | 92 *(59, 120)* | 86 *(71, 100)* | 52.5 | .17 |
| Anxiety Raw score | 17.0 *(0, 46)* | 14.5 *(4, 33)* | 78.0 | .94 |
| Internalising Raw score | 8.0 *(3, 32)* | 8.5 *(3, 26)* | 75.0 | .46 |
| Externalising Raw score | 9.0 *(1, 29)* | 9.0 *(0, 20)* | 79.0 | .58 |
| SCDC Total score | 7.0 *(0, 19)* | 4.0 *(0, 22)* | 75.0 | .46 |
| Inattention Raw score | 5.0 *(0, 14)* | 8.0 *(2, 11)* | 115.0 | .32 |
| Hyperactivity Raw score | 3.0 *(0, 14)* | 3.5 *(0, 10)* | 132.0 | .88 |

^a^ ‘Completer’ is a participant completing ≥ 4 blocks in Extinction phase; ‘Non-completer’ is a participant completing < 4 Extinction blocks.

^b^ Dp140- group includes Dp140-/71-

^c^ For FSIQ in non-completers n=7.

^d^ Significance (Sig.) testing for group comparisons conducted with Mann-Whitney U-test, taking alpha level *P*=.05.
