## Supplemental Table 6 for "Startle responses in Duchenne muscular dystrophy: a novel biomarker of brain dystrophin deficiency"

**Supplemental Table 6. Sensitivity analyses of primary outcomes, accounting for (A) non-completers, and (B) DMD participants taking B-blockers.**

| **A Sensitivity analysis including/excluding non-completers**^a^ | | | | | | | | | |
| --- | --- | --- | --- | --- | --- | --- | --- | --- | --- |
|  | **Mean differences *(95% Confidence Intervals)* for between groups comparisons**^b^  **(DMD vs. Control)** | | | | | | | | |
|  | **n** | **SCR_UC_*, µS***^c^ | **Sig., *P*** | **SCR_CS+_ ACQ1 block*, µS***^d^ | **Sig., *P*** | **SCR_EXT,_**  ***µS***^e^ | **Sig., *P*** | **ΔHR_UC_*, bpm***^f^ | **Sig., *P*** |
| DMD all | 28 | 3.1  *(1.0, 5.3)* | **.005** | 2.3  *(1.0, 3.6)* | **.001** | 0.37  *(-0.23, 0.96)* | 0.2 | -8.8  (-17.2, -0.3) | **.04** |
| DMD excl. non-completers | 20 | 3.5  *(1.3, 5.6)* | **.002** | 2.6  *(1.2, 3.9)* | **<.001** | 0.31  *(-0.3, 0.9)* | 0.30 | -9.5  (-18.4, -0.6) | **.04** |
| **B Sensitivity analysis including/excluding B-blocker participant**^b^ | | | | | | | | | |
|  |  | **ΔHR_UC_ within groups, in bpm** | | | | **Mean difference in ΔHR_UC_ *(95% Confidence Intervals),* in bpm, between DMD vs. Control** | | | |
|  | n | Mean ΔHR_UC_ *(sd)* | | Sig., *P,*  within group | | Mean diff. ΔHR_UC_ *(95%CI)* | | Sig., *P*  (DMD vs. Control) | |
| Control | 25 | -0.8 *(12.7)* | | .75 | |  | | | |
| DMD all | 31 | -6.4 *(9.7)* | | **.001** | | -8.7 *(-17.0, -0.5)* | | **.04** | |
| DMD excl. B-blockers | 30 | -6.6 *(9.9)* | | **.001** | | -9.1 *(-17.5, -0.7)* | | **.03** | |

^a^ Non-completers were defined as participants completing <4 extinction blocks. All primary SCR and HR metrics were derived prior to this time point.

^b^ Significance (Sig.) testing using univariate ANOVA adjusted for IQ for between-groups analyses, and repeated measures ANOVA for within-group analyses. Alpha level *P*=.05. *P*-values <.05 are highlighted in bold. Bonferroni adjustment made for multiple comparisons.

^c^ SCR_UC_ = Unconditioned skin conductance response to the first ‘threat’ trial with conditioned stimulus, CS+, in microSiemens (µS).

^d^ SCR_CS+_ ACQ1 = Skin conductance response to ‘threat’ CS+ trials in first Acquisition block.

^e^ SCR_EXT_ = Conditioned skin conductance response to the first CS+ trial of the Extinction phase, in µS.

^f^ ΔHR_UC_ = Unconditioned change in heart rate response to the first CS+ ‘threat’ trial, in beats per minute (bpm).
